# Impact of respiratory syncytial virus immunization in older adults: the importance of a valid burden estimation

**DOI:** 10.1101/2025.10.30.25339171

**Authors:** Charles-Antoine Guay, Radhouene Doggui, Patrick Fortin, Élise Fortin, Rachid Amini, Amandine Bemmo, Rodica Gilca

**Affiliations:** Centre de Recherche de l’Institut Universitaire de Cardiologie et de Pneumologie de Québec, Québec City, Québec, Canada; Département de médecine sociale et préventive, Faculté de médecine, Université Laval, Québec City, Québec, Canada; Direction des risques biologiques. Institut national de santé publique du Québec, Québec City, Québec, Canada; Département de microbiologie, infectiologie et immunologie, Faculté de médecine, Université de Montréal, Montréal, QC, Canada; Axe Maladies infectieuses et immunitaires, Centre de recherche du Centre hospitalier universitaire de Québec-Université Laval, Québec, QC, Canada

**Keywords:** Older adults, RSV, diagnostic test performance, vaccination, burden

## Abstract

**Introduction:** In the province of Québec, Canada, a respiratory syncytial virus (RSV) immunization program restricted to seniors residing in long-term care facilities (LTCF) and those aged ≥75 years from private senior residences (PSR) was implemented in Fall 2024, with approximately 65% vaccine uptake in the targeted populations. Despite high vaccine effectiveness, the real-world impact of the RSV immunization program on population-level averted hospitalizations and the influence that RSV diagnostic test performance might have remains unknown. This study aimed to evaluate the impact of the first season (2024-25) of the intervention among older adults in Québec while accounting for laboratory test performance.

**Methods:** A prospective hospital-based surveillance (HospiVir) and health administrative databases were used to assess the intervention impact. Observed to expected approach and before/after ecological design using a difference-in-difference Poisson regression, both accounting for seasonal variations and trends in comparison groups, were used. Hospitalization incidence rate estimates based on hospital-based surveillance were adjusted for the laboratory test performances using Lash equations.

**Results:** Based on HospiVir, a 49% (95% confidence interval (CI): -80– -5) decrease in RSV-associated hospitalizations was observed among adults aged ≥75 years residing in PSR. This decrease was 23% (95% CI: -38–66) in the overall population ≥75 years. A smaller decrease for the overall population ≥75 years was observed using health administrative data, but it did not reach statistical significance (10%; 95% CI: -7–23). The impact of the immunization program did not change following adjustment for diagnostic test performance, while the absolute burden of RSV-associated hospitalizations varied according to the proportion of RSV test positivity.

**Conclusions:** The 2024-25 RSV immunization program was associated with an almost 50% reduction in hospitalizations in residents of PSR aged ≥75 years. The magnitude of this reduction was roughly twofold lower when considering the overall population aged ≥75 years. Expanding the target population and strengthening vaccine promotion efforts to improve uptake are both essential to enhance the population-level impact of the RSV immunization program.

## 1. INTRODUCTION

Respiratory syncytial virus (RSV) is a major cause of respiratory-related morbidity and mortality worldwide, disproportionally affecting infants and older adults[1,2]. In high income countries, based on prospective studies (with laboratory-confirmed infection for RSV) the annual incidence of hospitalization ranges within 60-710 per 100,000 persons-year among those 75 years or older[3].

Two new RSV vaccines were introduced in Canada following their authorization for adults aged 60 years and older by Health Canada (between August 4^th^, 2023 and January 4^th^, 2024)[3]. In Québec, the second most populated province in Canada, after consideration of the Québec National Immunization Advisory Committee (*Comité sur l’immunisation du Québec*)[3], the Québec government decided to offer vaccination for the 2024-25 season to adults aged 60 years or more living in long-term care facilities (LTCF) as well as the most vulnerable of those aged 75 years and older living in private seniors’ residences (PSR)[4]. The vaccines were available on the private market for adults aged 60 years or more, but uptake was low ( ≈1% as of July 8^th^, 2025)[4]. By the end of the first season, vaccine coverage was around 10% in the general population aged 75 years and older; around 25% among all PSR residents (≈15% of the 75 years and older population), over 60% among the most vulnerable PSR residents, and around 65% in LTCF (<5% of the overall 75 years and older population)[4]. Increasing evidence from randomized controlled trials[5,6] and real-world studies[7–10] have shown a significant effectiveness of nearly 75-90% of RSV vaccines in preventing severe disease in older adults during the first season. However, the real-world impact of the RSV immunization program on population-level averted hospitalizations remains largely unknown.

One of the challenges of estimating the populational impact of RSV immunization programs in older adults are the difficulties in measuring the RSV burden. Indeed, questions have been raised concerning the lack of sensitivity of diagnostic approaches used in confirmation of the RSV diagnosis particularly[11–13]. Several preanalytical factors can impact sensitivity performances, irrespective of the used assay for RSV detection, such as the sampling day following the onset of symptoms (increasing delay associated with decreasing sensitivity), and the used sampling methods[13]. Simple multiplier methods have been proposed to correct for RSV burden under-ascertainments[11–13]. However, none of these accounted for the lack of specificity associated with diagnostic tools such as rapid antigen tests and multiplex PCRs[11–13]. Finally, none of the studies evaluated the extent to which diagnostic test performance characteristics specifically affect the assessment of the RSV vaccine program’s impact on the RSV burden.

Using data from both a prospective hospital-based surveillance and health administrative databases, we aimed to assess the impact of the 2024–25 RSV immunization program on RSV-associated hospitalizations among older adults in Québec, Canada and to determine whether laboratory test performance influences estimates of RSV hospitalization burden before and after program implementation. Particular attention was given to the number needed to vaccinate (NNV) and the number of avertable hospitalizations based on the pre-program RSV hospitalization burden to assess the theoretical impact of the intervention given the low vaccine uptake and the restricted targeted population during the first season of the program.

## 2. Methods

### 2.1. Study sources and study design

We used an observational before/after ecological study to assess the impact the impact of the immunization program (i.e., the intervention) on the RSV-associated hospitalization rates during the 2024-2025 season. Data were obtained from HospiVir, a prospective hospital-based surveillance network, and health administrative databases.

#### Prospective hospital-based surveillance

HospiVir provided prospective, clinically and laboratory-confirmed information on hospitalizations. The description of the HospiVir methodology has been reported elsewhere[14,15]. Two community and two academic/tertiary acute-care regional hospitals were participating since 2012-13 (catchment area of nearly 10% of the Québec population) and three additional tertiary hospitals joined the HospiVir network in 2021-22 and 2023-24 (catchment area for the six hospitals with adult population increased to 15%)[15,16]. In these hospitals, all patients admitted with acute respiratory infection (ARI) have a systematic deep nasal swab tested by a multiplex PCR at a central laboratory; those admitted for at least 24 hours are eligible (see Supplementary Table 1). Before the pandemic (2012-13 to 2018-19), the surveillance was conducted during the period of high influenza activity (when proportion of positive influenza tests reported by Québec’s sentinel laboratory surveillance network was ≥15% for at least two consecutive weeks), then extended to the entire year during the pandemic period and beyond[15]. Our analysis included all ARI hospitalizations with a laboratory-confirmed RSV infection. Information on the residency of patients in PSR and LTCF was available only from 2022-23 to 2024-25. Data on demographic and clinical details, including comorbid conditions were collected from medical charts.

#### Health administrative databases

Information on all hospitalizations in the province were obtained from the preliminary transmissions of the province’s acute-care admissions’ administrative database, MED-ECHO (*Maintenance et exploitation des données pour l’étude de la clientèle hospitalière*). Data on RSV-positive tests (available from 2022-23) was extracted from the provincial laboratory database.

Demographic data were retrieved from the *Fichier d’identification des personnes assurées de la Régie de l’assurance-maladie du Québec.* The presence of comorbidities was ascertained by using the Québec Integrated Chronic Disease Surveillance System (QICDSS)[17].

### 2.2. Study population and periods

The study population included all adults aged 65 years and older. We assessed the impact of the RSV immunization program in population aged 75 years and older residing in PSR that were hospitalized in one of the participating hospitals of HospiVir. LTCF residents were not included in the analysis since the numbers of those admitted for an ARI were limited and transfer practices from LTCF to hospitals fluctuated over the study periods. Adults aged 75 years and older residing in PSR represent around 15% of the overall 874,619 adults aged 75 years and older in Quebec based on the 2021-22 census data[15,19]. We also assessed the impact of the RSV immunization program in the overall population aged 75 years and older. In order to decrease the dilution of the effect given the limited vaccine uptake at the populational level, under the hypothesis that individuals targeted by the campaign will mostly concentrate among older population with comorbidities, we also conducted sensitivity analyses restricted to patients aged 80 years or more and those aged 85 years. Comparison groups were: 1) adults aged 65-74 years and 2) adults aged 75 years and older from the community in the analysis of PSR residents, as these groups were not targeted by the 2024–25 RSV immunization campaign. The 2024-25 season corresponds to the intervention period, while the pre-intervention period was defined according to the data availability (Supplementary Figure 1) and divided in pre-pandemic pre intervention period (2012-13 to 2018-19) and post-pandemic pre-intervention period (2022-23 to 2023-24).

### 2.3. Outcomes

The primary outcome was RSV-associated hospitalizations defined as a hospitalization with a RSV positive test at admission (i.e., in HospiVir) or 7 days prior to 3 days after admission (i.e., in MED-ECHO). In HospiVir, we estimated RSV-associated hospitalization rates stratified by age group, comorbidity and season, with adjustment for incomplete information, by using a similar approach to that used for estimation of influenza-associated hospitalizations[16]. To address potential misclassification of RSV infection due to imperfect sensitivity and specificity of multiplex PCR assays, we constructed a quantitative bias analysis model based on Lash’s equation for bias analysis[20] and the pooled estimates for sensitivity and specificity from a recent retrieved meta-analysis[13] (Supplementary Tables 2 and 3). Season-, age-, and comorbidity-specific correction factors were subsequently derived and applied to adjust for both under-and over-ascertainment of RSV-associated hospitalizations (Supplementary Table 4). Other outcomes, including RSV-associated emergency departments (ED) visits and RSV-associated death were assessed in exploratory analyses using health administrative data (Supplementary Table 5).

### 2.4. Statistical analyses

#### 2.4.1. Impact of the intervention

##### Prospective hospital-based data

We compared the observed-to-expected RSV-associated hospitalizations in 2024–25 among both intervention and comparison groups in HospiVir. Expected hospitalizations were derived using the ratio of hospitalizations in the intervention groups to those in the comparison groups (75 years old or more *vs*. 65-74 years; or 75 years old or more living in PSR *vs*. 75 years old or more from community) from the pre-intervention period (2023-24), multiplied by the observed number during the 2024-25 season. The assumption underlying the estimation of the expected hospitalizations was that the ratio of targeted over comparison groups from pre-intervention period would remain unchanged during the 2024-25 season in the absence of the intervention. This approach is not influenced by inter-seasonal variations in RSV severity, as it relies on the relative contribution of the targeted groups over comparison groups, rather than on the absolute burden observed in the preceding seasons as in traditional before–after comparisons. The relative reduction was primarily expressed as a proportion (relative impact) and secondary as a difference (absolute impact). We also conducted a sensitivity analysis using two pre-intervention years (2022-23 and 2023-24) for expected values for the comparability with the health administrative data-based analysis. Because the information on PSR was not available in HospiVir for 2022-23, only analysis comparing 75 years and older to 65-74 years was performed, similar to the analysis using health administrative databases.

##### Health administrative data

A difference-in-differences statistical method was used to control for temporal changes affecting all groups equally and to isolate the impact of the intervention. The analysis compared changes in hospitalization rates and other secondary outcomes (Supplementary Table 5) between the pre- and post-intervention periods among individuals aged 75 years old or more (intervention group), relative to changes in individuals aged 65 to 74 years (comparison group). Poisson regression was used to model the incidence rate of each outcome across the targeted and comparison groups. Robust Poisson regression model using generalized estimating equations was used to assess the risk ratio of complications for RSV-associated ED visits, accounting for nesting of patients within-hospitals. Regression models included the pre- and post-intervention periods, the intervention and comparison groups, and their interaction, which allows for estimating the adjusted relative impact of the intervention. By inverting the rate ratio or the risk ratio of the interaction term (1/RR) and multiplying it by the observed number of occurrences in the intervention group during the intervention period, we were able to estimate the expected numbers, rates, and proportions if no intervention had occurred as counterfactual scenario.

#### 2.4.2. Potential Impact of the intervention

##### NNV and potential avertable burden calculation

To assess the theorical impact of the intervention, we estimated the NNV to prevent one hospitalization for each season before the implementation of the campaign as 1/(RSV hospitalization rate x vaccine effectiveness (VE)) based on adjusted incidence rate estimates between 2012-13 and 2018-19 using HospiVir. Finally, we also explored different scenarios assuming VE of 70%, 80% and 90% and vaccine coverage of 10 % (i.e., similar to the 2024-25 vaccine coverage in the overall 75 years and older population), 30%, 60% and 90%.

All analyses were conducted by using SAS statistical software (version 9.4; SAS Institute, Inc.) and R Studio (version 2024.12.1+563). All estimates are presented with their corresponding 95% confidence intervals (CIs). A bootstrapping (n=10,000) was applied for the estimation of the 95% CI only for hospital-based estimates.

### 2.5. Ethics

The analyses were conducted using fully de-identified data within the framework of a program evaluation mandated by the Québec Ministry of Health. This project was deemed exempt from research ethics review by the CHU de Québec Research Ethics Board (#2026-8268).

## 3. RESULTS

### 3.1. Study population

Characteristics of the study participants from HospiVir for each of the study periods (i.e., pre-pandemic pre intervention period, post-pandemic pre-intervention period and intervention period) are shown in Table 1. The overall proportion of RSV-associated hospitalizations among ARI hospitalizations in adults ranged from 4% to 13% according to the season during the pre-pandemic period (Supplementary Figure 2). This proportion was 3% in 2022-23, 4% in 2023-24, and 6% in 2024-25 (data not shown). During the eight pre-pandemic seasons (2012-13 to 2018-19), 312 RSV-associated hospitalizations occurred among adults (≥18 years) (Table 1), including about 55% aged 75 years or more and 25% 65-74 years old. Post-pandemic, a total of 217 and 174 RSV-associated hospitalizations occurred in adults in 2022-23 to 2023-24 and in 2024-25, respectively, with 62% of those aged 75 years or more in 2022-23 to 2023-24 and 52% in 2024-25. Of those, 48% were admitted from PSR in 2022-23 to 2023-24 and 30% in 2024-25.

**Table 1.**
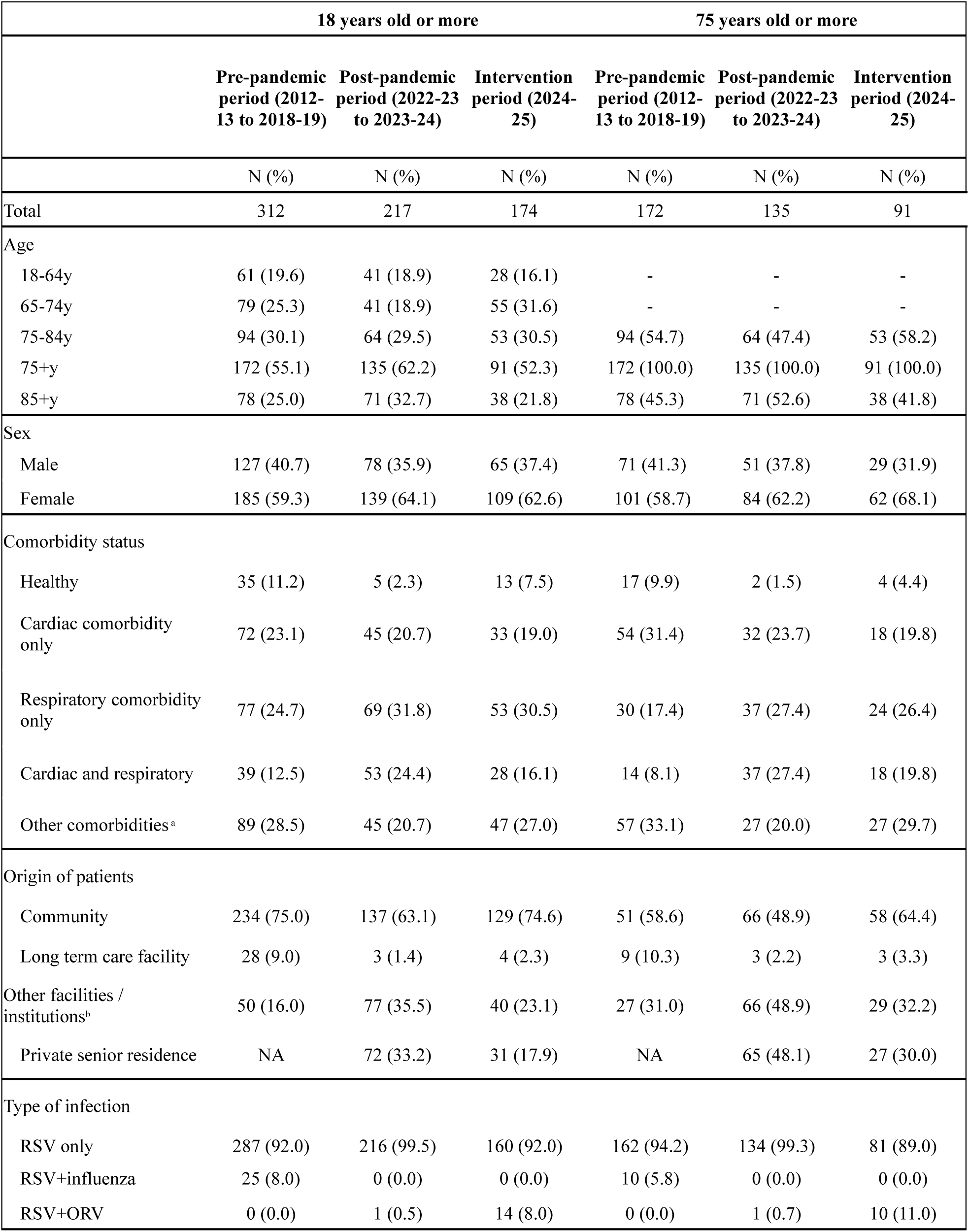
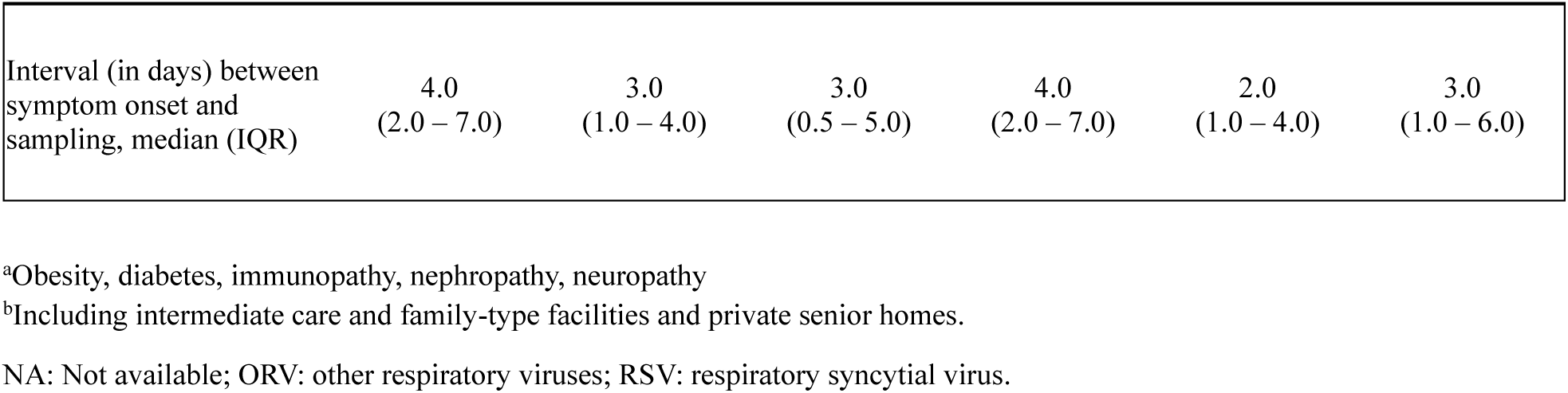
Characteristics of patients hospitalized with RSV in hospital-based prospective surveillance network in Québec, Canada according to study peridods

|  | 18 years old or more |  |  | 75 years old or more |  |  |
| --- | --- | --- | --- | --- | --- | --- |
|  | Pre-pandemic period (2012-13 to 2018-19) | Post-pandemic period (2022-23 to 2023-24) | Intervention period (2024-25) | Pre-pandemic period (2012-13 to 2018-19) | Post-pandemic period (2022-23 to 2023-24) | Intervention period (2024-25) |
|  | N (%) | N (%) | N (%) | N (%) | N (%) | N (%) |
| Total | 312 | 217 | 174 | 172 | 135 | 91 |
| Age |  |  |  |  |  |  |
| 18-64y | 61 (19.6) | 41 (18.9) | 28 (16.1) | - | - | - |
| 65-74y | 79 (25.3) | 41 (18.9) | 55 (31.6) | - | - | - |
| 75-84y | 94 (30.1) | 64 (29.5) | 53 (30.5) | 94 (54.7) | 64 (47.4) | 53 (58.2) |
| 75+y | 172 (55.1) | 135 (62.2) | 91 (52.3) | 172 (100.0) | 135 (100.0) | 91 (100.0) |
| 85+y | 78 (25.0) | 71 (32.7) | 38 (21.8) | 78 (45.3) | 71 (52.6) | 38 (41.8) |
| Sex |  |  |  |  |  |  |
| Male | 127 (40.7) | 78 (35.9) | 65 (37.4) | 71 (41.3) | 51 (37.8) | 29 (31.9) |
| Female | 185 (59.3) | 139 (64.1) | 109 (62.6) | 101 (58.7) | 84 (62.2) | 62 (68.1) |
| Comorbidity status |  |  |  |  |  |  |
| Healthy | 35 (11.2) | 5 (2.3) | 13 (7.5) | 17 (9.9) | 2 (1.5) | 4 (4.4) |
| Cardiac comorbidity only | 72 (23.1) | 45 (20.7) | 33 (19.0) | 54 (31.4) | 32 (23.7) | 18 (19.8) |
| Respiratory comorbidity only | 77 (24.7) | 69 (31.8) | 53 (30.5) | 30 (17.4) | 37 (27.4) | 24 (26.4) |
| Cardiac and respiratory | 39 (12.5) | 53 (24.4) | 28 (16.1) | 14 (8.1) | 37 (27.4) | 18 (19.8) |
| Other comorbidities<sup>a</sup> | 89 (28.5) | 45 (20.7) | 47 (27.0) | 57 (33.1) | 27 (20.0) | 27 (29.7) |
| Origin of patients |  |  |  |  |  |  |
| Community | 234 (75.0) | 137 (63.1) | 129 (74.6) | 51 (58.6) | 66 (48.9) | 58 (64.4) |
| Long term care facility | 28 (9.0) | 3 (1.4) | 4 (2.3) | 9 (10.3) | 3 (2.2) | 3 (3.3) |
| Other facilities / institutions<sup>b</sup> | 50 (16.0) | 77 (35.5) | 40 (23.1) | 27 (31.0) | 66 (48.9) | 29 (32.2) |
| Private senior residence | NA | 72 (33.2) | 31 (17.9) | NA | 65 (48.1) | 27 (30.0) |
| Type of infection |  |  |  |  |  |  |
| RSV only | 287 (92.0) | 216 (99.5) | 160 (92.0) | 162 (94.2) | 134 (99.3) | 81 (89.0) |
| RSV+influenza | 25 (8.0) | 0 (0.0) | 0 (0.0) | 10 (5.8) | 0 (0.0) | 0 (0.0) |
| RSV+ORV | 0 (0.0) | 1 (0.5) | 14 (8.0) | 0 (0.0) | 1 (0.7) | 10 (11.0) |
| Interval (in days) between symptom onset and sampling, median (IQR) | 4.0<br>(2.0 – 7.0) | 3.0<br>(1.0 – 4.0) | 3.0<br>(0.5 – 5.0) | 4.0<br>(2.0 – 7.0) | 2.0<br>(1.0 – 4.0) | 3.0<br>(1.0 – 6.0) |

<sup>a</sup>Obesity, diabetes, immunopathy, nephropathy, neuropathy
<sup>b</sup>Including intermediate care and family-type facilities and private senior homes.
NA: Not available; ORV: other respiratory viruses; RSV: respiratory syncytial virus.

### 3.2. Adjustment for diagnostic test performance

We used the eight pre-pandemic RSV seasons (2012-13 to 2018-19) to develop a model that incorporates diagnostic test performance in estimating the burden of RSV-associated hospitalizations (Table 2). As expected, the adjustment of incidence rate for laboratory test sensitivity resulted in an increase of about 10% with the same magnitude across all age groups, with an overall incidence of 122.0 per 100 000 person-years in those aged 75 years or more (Table 2). Conversely, the adjusted incidence rate for laboratory test specificity decreased by about 10 % the overall incidence rate. After adjustment for both sensitivity and specificity, estimated hospitalization incidence rates ranged from a reduction of 10% to an increase of approximately 5% compared to the unadjusted ones, depending on the season (Table 2 and Supplementary Table 6). Adjustment showed comparable seasonal variations regardless of age groups and comorbid status. In average, adjustment for specificity and sensitivity resulted in lower estimated hospitalization rates compared to unadjusted rates in seasons where RSV positivity proportion (i.e., the proportion of hospitalizations with a positive RSV test in HospiVir) was below 10% (Supplementary Table 6). Adjusted rates tended to be higher in seasons where RSV prevalence exceeded 10%.

**Table 2.** RSV-associated hospitalization rates in hospital-based prospective surveillance network according to age, comorbidity status and adjustment method between 2012-2013 and 2018-2019

|  | **Healthy** |  | **Comorbidity** |  | **Overall** |  |
| --- | --- | --- | --- | --- | --- | --- |
| Age groups | Adjusted RSV hospitalization rate per 100 000 (95% CI) | Rate difference (%) | Adjusted RSV hospitalization rate per 100 000 (95% CI) | Rate difference (%) | Adjusted RSV hospitalization rate per 100 000 (95% CI) | Rate difference (%) |
| **Without adjustment for diagnostic test performance** |  |  |  |  |  |  |
| 18-64 years | 1.6 (1.0 – 2.1) | NA | 10.3 (8.4 – 12.2) | NA | 4.6 (3.8 – 5.3) | NA |
| 65-74 years | 3.2 (1.1 – 5.4) | NA | 82.8 (70.6 – 95.0) | NA | 39.1 (33.4 – 44.7) | NA |
| 75-84 years | 19.6 (11.5 – 27.6) | NA | 141.8 (123.2 – 160.4) | NA | 89.7 (78.5 – 101.0) | NA |
| 75+ years | 26.7 (18.6 – 34.7) | NA | 171.9 (154.8 – 188.9) | NA | 112.0 (101.4 – 122.6) | NA |
| 85+ years | 46.1 (25.7 – 66.6) | NA | 239.7 (203.3 -271.1) | NA | 166.4 (142.5 -190.3) | NA |
| Overall | 2.5 (1.9 -3.0) | NA | 45.2 (41.8 – 48.6) | NA | 16.7 (15.5 -17.9) | NA |
| **Adjustment for diagnostic test sensitivity** |  |  |  |  |  |  |
| 18-64 years | 1.7 (1.1 – 2.3) | 6.3 | 11.2 (9.2 – 13.3) | 8.7 | 5.0 (4.2 – 5.8) | 8.7 |
| 65-74 years | 3.5 (1.3 – 5.8) | 9.4 | 90.3 (77.6 – 103.0) | 9.1 | 42.6 (36.7 – 48.4) | 9.0 |
| 75-84 years | 21.1 (12.7 – 29.4) | 8.2 | 154.6 (135.1 -174.0) | 9.0 | 97.7 (86.0 – 109.4) | 8.9 |
| 75+ years | 28.9 (20.5 – 37.2) | 7.7 | 187.3 (169.5.1 – 205.2) | 9.0 | 122.0 (111.0 – 133.0) | 8.9 |
| 85+ years | 50.3 (29.0 – 71.6) | 9.1 | 261.3 (223.3 – 299.3) | 9.0 | 181.3 (156.4 -206.3) | 9.0 |
| Overall | 3.0 (2.4 – 3.6) | 16.7 | 49.2 (45.7 – 52.8) | 8.8 | 18.4 (17.1 – 19.7) | 10.2 |
| **Adjustment for diagnostic test specificity** |  |  |  |  |  |  |
| 18-64 years | 1.3 (0.8 – 1.8) | -18.8 | 9.1 (7.3 – 10.9) | -11.6 | 4.0 (3.3 – 4.7) | -13.0 |
| 65-74 years | 2.8 (0.8 – 4.8) | -12.5 | 73.4 (62.0 – 84.9) | -11.4 | 34.6 (29.3 – 39.9) | -11.5 |
| 75-84 years | 17.2 (9.7 – 24.8) | -10.9 | 124.7 (107.3 – 142.2) | -12.1 | 78.9 (68.4 – 89.5) | -12.0 |
| 75+ years | 23.8 (16.2 – 31.4) | -10.3 | 153.3 (137.2 – 169.5) | -10.8 | 99.9 (90.0 – 109.9) | -10.8 |
| 85+ years | 42.0 (22.5 – 61.5) | -8.9 | 217.9 (183.2 – 252.6) | -9.1 | 151.2 (128.5 – 174.0) | -9.1 |
| Overall | 2.4 (1.9 – 3.0) | -5.6 | 40.2 (37.0 – 43.4) | -11.1 | 15.0 (13.9 - 16.1) | -10.2 |
| **Adjustment for diagnostic test sensitivity and specificity** |  |  |  |  |  |  |
| 18-64 years | 1.5 (0.9 – 2.0) | -6.3 | 10.0 (8.0 – 11.9) | -2.9 | 4.4 (3.6 – 5.1) | -4.3 |
| 65-74 years | 3.1 (1.0 – 5.3) | -3.1 | 80.4 (68.4 – 92.4) | -2.9 | 37.9 (32.4 – 43.4) | -3.1 |
| 75-84 years | 18.9 (11.0 – 26.8) | -3.6 | 136.7 (118.4 – 155.0) | -3.6 | 86.5 (75.5 -97.6) | -3.6 |
| 75+ years | 26.1 (18.2 – 34.1) | -2.2 | 167.9 (151.0 -184.7) | -2.3 | 109.4 (99.0 – 119.9) | -2.3 |
| 85+ years | 46.0 (25.6 -66.4) | -0.2 | 238.1 (201.8 – 274.4) | -0.7 | 165.3 (141.5 -189.1) | -0.7 |
| Overall | 2.7 (2.1 – 3.2) | 8.0 | 44.0 (40.6 – 47.3) | -2.7 | 16.4 (15.2 -17.6) | -1.8 |

RSV: respiratory syncytial virus; CI: confidence interval; NA: not applicable

### 3.3. Impact of the intervention

#### Based on the hospital-based prospective surveillance network

Using data from HospiVir, RSV-associated hospitalizations were reduced by 49% (95% CI: -80-5) in adults aged 75 years or more living in PSR compared to the expected number of hospitalizations based on the 65-74 years old comparison group (Table 3). This decrease was 48% (95% CI: -69 – -26) when compared to the adults aged 75 years or more living in the community. The overall reduction of RSV-associated hospitalizations in adults aged 75 years or more was 23% (95% CI: -66 – 38) using those aged 65-74 years to compute the ratio used to estimate the expected hospitalizations. Adjustment for the laboratory test performance did not change the estimated relative decrease but the absolute difference and the number of prevented hospitalizations were numerically lower (Table 3). Point estimated of relative and absolute impact of the vaccination campaign tended to increase in older age groups, where the target population is expected to be more prevalent. Finally, similar associations were observed when using the 2022-23 and 2023-24 as comparator period (Supplementary Table 7).

#### Based on health administrative data: Difference-in-difference analyses

##### RSV-associated ED visits, hospitalizations and deaths

Figure 1 shows the weekly incidence (100 000 person-years) of hospitalizations derived from health administrative databases during the study period (2022-23 to 2024-25 seasons) in adults aged 65-74 years and those aged 75 years or more. Visually, the difference between the two age groups was consistent during the two examined seasons before the intervention. The reduction in the risk of RSV-associated hospitalization in those 75 years old or more, taking into account time trends in those 65-74 years old, was of 10% (95% CI: -7 – 23), without reaching statistical significance when (Table 4). Moreover, in the analysis restricted to those aged 85 years or more, the relative risk reduction of RSV-associated hospitalization (21%; 95% CI: -34 – -4) was greater and statistically significant. The reduction in the risk of RSV-associated ED visits (from 10% to 25 %) and RSV-associated death (34% to 47%) showed the same trends of increasing magnitude of point estimates with age, with statistically significant estimates for those aged 80 years or more (Supplementary figure 3, Supplementary Tables 8 and 9).

**Figure 1.**
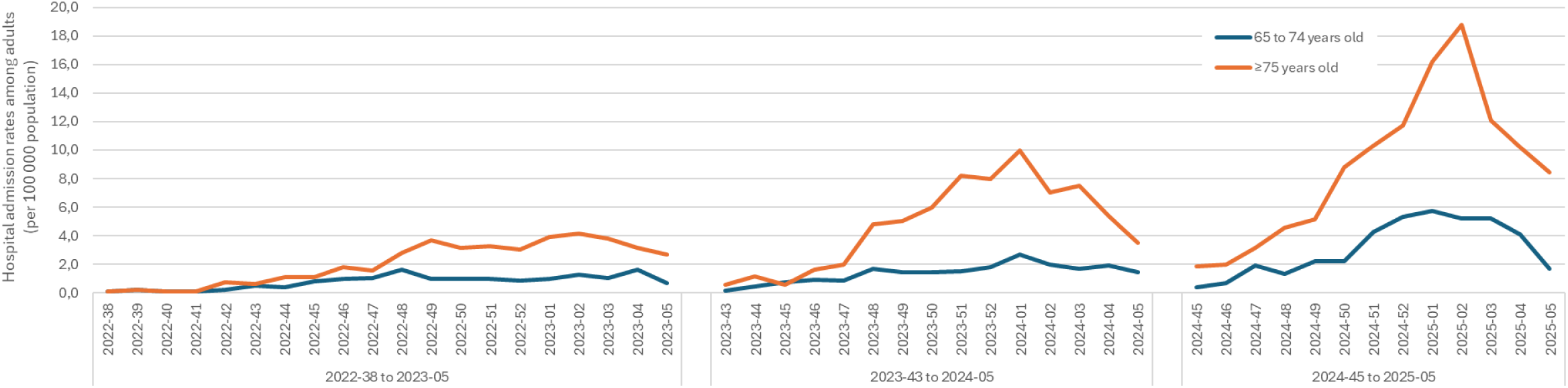
Weekly incidence of RSV-associated hospitalizations based on the health administrative data in Québec, Canada (2022-2023 to 2024-2025).

**Table 3.** Observed and expected RSV-associated hospitalizations in hospital-based prospective surveillance network following the RSV immunization campaign in adults aged 75 years and older in Québec, Canada, **using 1 pasts season (2023-24) as reference**

| Burden estimation method | Targeted group | Observed RSV-associated hospitalizations, n | Expected RSV-associated hospitalizations, n (95% CI) | Absolute impact measure, n (95 % CI) | Relative impact measure, % (95 %CI) | P value<sup>a</sup> (Exact Poisson) |
| --- | --- | --- | --- | --- | --- | --- |
| Without adjustment for diagnostic test performance | Group used to estimate the difference or ratio for the targeted group**: Individuals aged 65-74 years<br>N<sub>2023-24</sub> = 19; N<sub>2024-25</sub> = 34 |  |  |  |  |  |
|  | 75+ years living in PSR | 20 | 39 (21 – 98) | -19 (-78 – -1) | -49 (-80 – -5) | 0.001 |
|  | All 75+ years | 66 | 86 (48 – 195) | -20 (-129 – 18) | -23 (-66 – 38) | 0.03 |
|  | 80+ years living in PSR | 17 | 34 (19 – 44) | -17 (-30 – -2) | -50 (-61 – -9) | 0.002 |
|  | All 80+ years | 49 | 66 (43 – 75) | -17 (-26 – 6) | -26 (-34 – 15) | 0.04 |
|  | 85+ years living in PSR | 14 | 29 (14 – 41) | -15 (-27 – 0) | -51 (-66 – 0) | 0.004 |
|  | All 85+ years | 30 | 41 (25 – 49) | -11 (-19 – 5) | -27 (-39 – 20) | 0.09 |
| Adjustment for diagnostic test sensitivity and specificity | Group used to estimate the difference or ratio for the targeted group**: Individuals aged 65-74 years<br>N<sub>2023-24</sub> = 16.53; N<sub>2024-25</sub> = 31.96 |  |  |  |  |  |
|  | 75+ years living in PSR | 19 | 37 (20 – 92) | -18 (-73 – -1) | -49 (-80 – -5) | 0.003 |
|  | All 75+ years | 62 | 81 (45 – 184) | -19 (-122 – 17) | -23 (-66 – 38) | 0.06 |
|  | 80+ years living in PSR | 16 | 32 (17 – 41) | -16 (-25 – -1) | -50 (-61 – -9) | 0.003 |
|  | All 80+ years | 46 | 62 (40 – 70) | -16 (-24 – 6) | -26 (-34 – 15) | 0.07 |
|  | 85+ years living in PSR | 13 | 27 (13 – 38) | -14 (-25 – 0) | -51 (-66 – 0) | 0.006 |
|  | All 85+ years | 28 | 39 (23 – 46) | -11 (-18 – 5) | -27 (-39 – 20) | 0.12 |
|  | Group used to estimate the ratio for the targeted group**: Individuals aged 75+ years living in community<br>N<sub>2023-24</sub> = 24; N<sub>2024-25</sub> = 42 |  |  |  |  |  |

| **Burden estimation method** | **Targeted group** | **Observed RSV-associated hospitalizations, n** | **Expected RSV-associated hospitalizations, n (95% CI)** | **Absolute impact measure, n (95 % CI)** | **Relative impact measure, % (95 %CI)** | **P value<sup>a</sup> (Exact Poisson)** |
| --- | --- | --- | --- | --- | --- | --- |
| Without adjustment for diagnostic test performance | 75+ years living in PSR | 20 | 39 (27 – 65) | -19 (-45 – -7) | -48 (-69 – -26) | 0.002 |
| Adjustment for diagnostic test sensitivity and specificity | Group used to estimate the difference or ratio for the targeted group<sup>**</sup>: Individuals aged 75+ years living in community<br>N<sub>2023-24</sub> = 20.88; N<sub>2024-25</sub> = 39.48 |  |  |  |  |  |
|  | 75+ years living in PSR | 19 | 36 (26 – 61) | -17 (-42 – -7) | -48 (-69 – -26) | 0.004 |

<sup>a</sup>P values calculated using an exact statistical test based on the Poisson distribution. PSR: private senior homes; RSV: respiratory syncytial virus; CI: confidence interval

**Table 4.** Changes in observed number of RSV associated outcomes among older adults in 2024-25 compared to the expected number in counterfactual scenario without the immunization campaign (2022-2023 to 2023-24), based on health administrative data in Québec, Canada

| Outcome | Reduction adjusted for time trends in younger adults (%<br>95% CI) | Rates or proportions post-intervention |  | Number of severe cases post-intervention |  | Number of prevented cases (95% CI) |
| --- | --- | --- | --- | --- | --- | --- |
|  |  | Observed | Expected (95% CI) | Observed | Expected (95% CI) |  |
| Hospital admissions with a positive RSV test - $\geq 75$ years old (rate per 100 000) | 10% (-7 – 23) | 113.35 | 125.33<br>(106.24 – 147.84) | 966 | 1068 (905 – 1260) | 102 admissions<br>(-61 – 294) |
| Hospital admissions with a positive RSV test - $\geq 80$ years old with comorbidities (rate per 100 000) | 15% (-1 – 29) | 159.99 | 188.05<br>(157.94 – 223.89) | 670 | 787 (661 – 938) | 117 admissions<br>(-9 – 268) |
| Hospital admissions with a positive RSV test - $\geq 85$ years old with comorbidities (rate per 100 000) | 21% (4 – 34) | 192.67 | 242.57<br>(200.20 – 293.89) | 411 | 517 (427 – 627) | 106 admissions<br>(16 – 216) |

RSV: respiratory syncytial virus

### 3.4. NNV and avertable hospitalizations

Table 5 presents the NNVs and the estimated number of RSV-associated hospitalizations that could be averted by age group, based on pre-pandemic RSV rates from HospiVir, according to different VE and vaccine coverage scenarios (i.e., the theoretical impact of the immunization program). In average, the NNVs are 38-fold higher (32,628 vs 864) in adults aged 18-64 years compared to adults of 85 years and older (assuming a vaccine effectiveness of 70%). NNVs are lower in patients with comorbidities in all groups. In individuals aged 75 years and older, between 47 (VE=70%, coverage =10%) and 544 (VE=90%, coverage 90%) could be averted, with the highest number of averted hospitalizations observed among individuals with underlying comorbidities. Results from sensitivity analyses exploring different VE and vaccine coverage scenarios for each pre-pandemic seasons before the intervention are presented in Supplementary Table 10.

**Table 5.** Number needed to vaccinate to prevent an RSV hospitalization and avertable hospitalizations according to age groups and comorbidity status between 2012-2013 and 2018-2019, Québec, Canada.

|  | Age groups | RSV hospitalization rate per 100,000 | Vaccine effectiveness (%) | Number needed vaccinate | Avertable hospitalizations |  |  |  |
| --- | --- | --- | --- | --- | --- | --- | --- | --- |
|  |  |  |  |  | VC=10% | VC=30% | VC=60% | VC=90% |
| All patients | 18-64 y | 4.4 | 70 | 32,628 | 16 | 48 | 96 | 144 |
|  |  |  | 80 | 28,550 | 18 | 55 | 110 | 165 |
|  |  |  | 90 | 25,378 | 21 | 62 | 124 | 186 |
|  | 65-74 y | 37.9 | 70 | 3,769 | 22 | 65 | 131 | 196 |
|  |  |  | 80 | 3,298 | 25 | 75 | 149 | 224 |
|  |  |  | 90 | 2,932 | 28 | 84 | 168 | 252 |
|  | 75-84 y | 86.5 | 70 | 1,651 | 26 | 79 | 157 | 236 |
|  |  |  | 80 | 1,444 | 30 | 90 | 180 | 269 |
|  |  |  | 90 | 1,284 | 34 | 101 | 202 | 303 |
|  | 75+y | 109.4 | 70 | 1,306 | 47 | 141 | 282 | 423 |
|  |  |  | 80 | 1,142 | 54 | 161 | 322 | 484 |
|  |  |  | 90 | 1,015 | 60 | 181 | 363 | 544 |
|  | 85+ y | 165.3 | 70 | 864 | 21 | 63 | 126 | 189 |
|  |  |  | 80 | 756 | 24 | 72 | 144 | 216 |
|  |  |  | 90 | 672 | 27 | 81 | 162 | 243 |
| Patients with comorbidity | 18-64 y | 10.0 | 70 | 14,356 | 12 | 37 | 75 | 112 |
|  |  |  | 80 | 12,562 | 14 | 43 | 86 | 129 |
|  |  |  | 90 | 11,166 | 16 | 48 | 96 | 145 |
|  | 65-74 y | 80.4 | 70 | 1,777 | 21 | 62 | 125 | 187 |
|  |  |  | 80 | 1,555 | 24 | 71 | 143 | 214 |
|  |  |  | 90 | 1,382 | 27 | 80 | 161 | 241 |
|  | 75-84 y | 136.7 | 70 | 1,045 | 24 | 71 | 142 | 214 |
|  |  |  | 80 | 914 | 27 | 81 | 163 | 244 |
|  |  |  | 90 | 813 | 31 | 92 | 183 | 275 |
|  | 75+y | 167.9 | 70 | 851 | 42 | 127 | 254 | 382 |
|  |  |  | 80 | 745 | 48 | 145 | 291 | 436 |
|  |  |  | 90 | 662 | 55 | 164 | 327 | 491 |
|  | 85+ y | 238.1 | 70 | 600 | 19 | 56 | 113 | 169 |
|  |  |  | 80 | 525 | 21 | 64 | 129 | 193 |
|  |  |  | 90 | 467 | 24 | 72 | 145 | 217 |

CI: confidence interval; RSV: respiratory syncytial virus; VC: vaccine coverage

## DISCUSSION

The study evaluated the impact of the immunization strategy on reducing the RSV-associated hospitalizations during the first season (2024-25) of the intervention in the province of Québec, Canada, by using both active prospective surveillance data and retrospective health administrative databases. We found the intervention to be associated with a reduction of RSV-associated hospitalizations in adults aged 75 years and older, adjusted for trends in comparison groups and seasons. Our findings indicate that the intervention had its most significant impact within the targeted group of residents in PSR (48%; 95% CI: -69 – -26), representing approximatively twice the impact estimated in the analysis of all adults aged 75 years and older, including only a proportion (around 40%) of those targeted by the immunization campaign. Furthermore, the impact detected using administrative databases including overall Quebec population of 75 years and older was numerically lower (10%; 95% CI: -7– 23) compared to - 23% (95% CI: -66 – 38) based on HospiVir. This may indicate differences in the representation of PSR residents and other population characteristics between the overall population and those captured within a sentinel surveillance network such as HospiVir. We also observed important variations in the avertable RSV-associated hospitalizations according to the age groups and vaccine coverage. These observations suggest that the number of averted hospitalizations by the RSV immunization program in older adults would increase with broader target populations and higher vaccine uptake.

A VE of at least 75% against RSV-associated hospitalization was reported during the first season following the vaccination campaign among older adults[6–9]. However, high VE estimates do not translate necessarily to a significant reduction in hospitalizations rates at the population level. Indeed, the population-level impact of an immunization program is determined not only by the individual-level effectiveness of the vaccine but it is also highly correlated to the extent of vaccine uptake[21,22]. In 2024-25, the publicly funded RSV immunization program in older adults was limited to the more vulnerable adults living in PSR and LTCF. The vaccine coverage reached around 25% (over 60% in the most vulnerable) in PSR and 65% in LTCF, by the end of the season, which led to an almost 50% reductions in RSV-associated hospitalizations in residents of PSR aged 75 years and older. Although the impact of the immunization program could not be specifically assessed in resident of LTCF, similar reduction, with similar vaccine uptake, should be expected as VE did not significantly differ in older age groups and in patients with more comorbidities[5–10]. Furthermore, although not always statistically significant, we found a trend toward a higher impact of the immunization program in older age groups both in HospiVir and health administrative data. These results are consistent with our primary analysis restricted to PSR residents since a greater proportion of older adults lives in PSR and LTCF as age increases. For instance, nearly 30% of Quebecers 85 years old or over live in PSR[23] and it is in this age group that the greatest impact of the immunization programme was observed. These findings illustrate that, even in the absence of information on place of residence (or other information on targeted population), health administrative data can be effectively used to approximate the impact of an immunization program in the overall population by concentrating on groups where the dilution of the effect would be minimal. However, such estimates are likely to remain conservative and are conditional on a minimal threshold of vaccine uptake allowing detectability We did not observe a significant decrease in RSV-associated hospitalizations in the overall population aged 75 years and older, which could be explained by the low vaccine coverage (≈10%) reached. In comparison, Hammed and al.[24] found a reduction of 62.1% (95% CI 35.0-79.8) in RSV-associated hospitalization (defined as an RSV positive test 14 days before or within 2 days of hospital admission) in adults aged 75 to 79 years following the implementation of RSV vaccines in the United Kingdom in 2025. The vaccine coverage in this study reached 68.6% in the targeted age group[24]. On the other hand, we found that a large number of persons aged 75 years and older may need to be vaccinated to prevent a substantial number of events (expressed as NNV), ranging between 864 and 1,142 according to the presumed VE and vaccine uptake. High NNV despite high VE can be explained by relatively low baseline burden of disease before the implementation of a vaccine strategy. Nonetheless, as shown in our study, disease burden and avertable hospitalizations increase substantially by age and comorbidity status. Maximizing the population-level impact of an RSV vaccination strategy, requires prioritizing vaccine promotion to achieve high coverage in groups experiencing the greatest disease burden. Even with the current targeted population (about 20% of the overall 75 years and older population), we found that the intervention had a detectable effect at the population level. Indeed, individuals 75 years and older in LTCF and PSR were found to have the highest burden of disease underscoring the importance of attaining high vaccination coverage within this priority population[25,26].

Accurately quantifying the impact of an immunization program requires a valid and precise estimation of the burden of the disease before and after the implementation of the program[31,32]. With the approval of new vaccines against RSV, many groups have tried to accurately quantify the burden of RSV in older adults to guide immunization recommendations. The main concern of these studies on burden of disease before the implementation of RSV immunization programs was the possible underascertainment of RSV cases associated with reduced sensitivity of RSV diagnostic tests in adults. McLaughlin et al.[12] were among the first to propose a simple multiplier approach to estimate the burden of RSV while adjusting for under-ascertainment related to incomplete testing and suboptimal test sensitivity. Their systematic review and meta-analysis of diagnostic methods for RSV yielded a median detection multiplier of 1.5×, which several public health organizations have subsequently adopted to adjust observed incidence rates in adults when quantifying RSV burden[12]. Subsequent studies have suggested alternative multipliers ranging from 1.5 to 3.5[11–13]. However, these straightforward adjustment approaches do not comprehensively account for the full spectrum of diagnostic test performance characteristics, including specificity and predictive values[11–13]. In our study, we explicitly considered the performance characteristics of the multiplex PCR assay employed in our model for burden estimation, taking into account both its sensitivity and specificity. Overall, we found between 4.3% and 0.7% lower incidence rates estimate for RSV-associated hospitalizations compared to the ones obtained without adjustment for diagnostic test performance, varying inversely with age groups and comorbidity status. Furthermore, the impact of the immunization program did not change following adjustment for diagnostic test performance, which is consistent with stable sensitivity and specificity values that are intrinsic measure of a test accuracy, independent of the prevalence[27,28]. However, our findings indicate that the effect of adjustment on the estimated burden varied with RSV positivity; incidence rates were notably underestimated when test performance was not accounted for, particularly in contexts where RSV positivity tended to exceed 10% and overestimated otherwise. Consequently, our findings highlight the importance of disease prevalence and positive predictive values when adjusting for diagnostic test performance.

Our study benefits from drawing on two complementary methodological approaches to provide a more comprehensive assessment of the impact of the intervention among older adults in Québec. The strengths of the primary analysis were prospective, active hospital-based comprehensive recruitment of patients with stringent criteria, coupled with systematic testing for RSV by using a multiplex PCR. These strengths most likely reduced the misclassification bias. The secondary analysis, based on the administrative health data, offers a better statistical power as it covers the entire population level. However, several limitations must be acknowledged.

In the prospective surveillance approach, the limited number of cases in the targeted group led to a wide confidence interval, precluding a definitive conclusion regarding the magnitude of the intervention. Some cases would have been missed because elderly may present with non typical respiratory symptoms, such as confusion, anorexia, worsening of congestive heart failure that may challenge the identification of the RSV associated hospitalizations[29]. The proportion of such missed cases is expected to be consistent across the study period. On the other hand, the health administrative data is subject to misclassification bias since no direct evaluation of the patients is made[30]. In addition, it could be impacted by variations in testing practices over time and the propensity to test (increasing with time, Supplementary Figure 3)[31], which is often associated with an underestimation of RSV burden[32]. The magnitude and direction of bias in program impact estimates are less predictable when using health administrative databases, as they depend on variations in clinical and testing practices before and after the implementation of the intervention that can lead to differential misclassification. However, the difference-in-difference method accounted for changes in testing practices over time under the assumption that these changes were similar in both targeted and comparison age groups. The use of health administrative data also presents challenges in accurately delineating the population targeted by the intervention. In contrast to our hospital-based surveillance approach, health administrative data did not allow us to identify individuals residing in PSR and we had to rely on an age-based definition, which may dilute the estimated impact of the intervention, given that most of individuals aged 75 years and older were not vaccinated.

Despite these limitations, we showed consistent decrease in RSV-associated hospitalizations in older adults by using different approaches, providing promising findings after the first year of the RSV vaccination campaign, which may serve as a foundation for strengthening vaccination promotion in high-priority vulnerable populations.

## CONCLUSION

This is the first study that assessed the real-world impact of the RSV immunization campaign using both hospital-based prospective and administrative data-based designs and two different statistical approaches. Results were consistent across methods used and showed the role of the vaccination in reducing the burden of disease associated to the RSV. The study also underlines the importance of higher vaccine uptake for achieving a more substantial effect on a populational level. More efforts are needed to increase the vaccine coverage, especially in vulnerable populations through vaccine promotion and identification of barriers. Furthermore, accurate estimation of the RSV burden is essential for an accurate assessment of the impact of the RSV vaccination. We showed that accounting for laboratory test performance could lead to a certain change in burden estimates, with a minimal influence on the evaluation of the program’s impact.

### Declarations

#### Ethics approval and consent to participate

This study do not contain any individual person’s data. This study was deemed exempt from research ethics review by the CHU de Québec Research Ethics Board (#2026-8268).

#### Consent for publication

Not applicable.

#### Availability of data and materials

The datasets generated and analysed during the study are not publicly available due to individual privacy stakes. The data are stored on a secure server and regulated by the Quebec Ministry of Health and the Quebec Health Insurance Board. Access to the data is restricted to authorized personnel only to prevent any sensitive data breaches. Any requests should be addressed to the Ministère de la Santé et des Services sociaux du Québec. Code can be shared upon request by the authors.

#### Author’s contribution

Guay and Doggui had full access to all of the data in the study and take responsibility for the integrity of the data and the accuracy of the data analysis. **Concept and design**: Guay, Doggui,

Gilca. **Acquisition, analysis, or interpretation of data**: All authors. **Drafting of the manuscript**: Guay, Doggui, Gilca. **Critical revision of the manuscript for important intellectual content**: All authors. **Statistical analysis**: Guay, Doggui, Fortin P, Fortin É, Amini, Bemmo. **Supervision**: Gilca.

## Data Availability

The datasets generated and analysed during the study are not publicly available due to individual privacy stakes. The data are stored on a secure server and regulated by the Quebec Ministry of Health and the Quebec Health Insurance Board. Access to the data is restricted to authorized personnel only to prevent any sensitive data breaches. Any requests should be addressed to the Ministere de la Sante et des Services sociaux du Quebec. Code can be shared upon request by the authors.

## Acknowledgments

We acknowledge France Bouchard who coordinated the HospiVir surveillance program, and Josiane Rivard for technical support. We are extremely grateful to the front-line surveillance staff from the participating hospitals who collected and provided data: Francois Ménard and team from Chicoutimi Hospital; Jeannot Dumaresq and team from Hôtel-Dieu de Lévis Hospital; Patrick Dolce and team from Rimouski Hospital; Alexis Danylo and team from Trois-Rivières Hospital; Charles Frenette (deceased as of September 23, 2025) and team from Centre universitaire de santé McGill. We also thank Lyne Desautels and Hugues Charest from Laboratoire de Santé Publique du Québec. Finally, we thank Sara Carazo for insightful discussions that helped refine the conceptual framework and study design.

## Footnotes

None.

## Supplementary Information

This article has an online supplementary material.

**Abbreviations list**
ARI: Acute respiratory infection
CI: Confidence interval
ED: Emergency departments
LTCF: Long-term care facilities
MED-ECHO: Maintenance and Use of Data for the Study of Hospital
Clientele NNV: Number needed to vaccinate
QICDSS: Quebec Integrated Chronic Disease
Surveillance System PSR: Private senior residences
RAMQ: Quebec Health Insurance Board
RR: Relative risk
RSV: Respiratory Syncytial Virus
VE: Vaccine effectiveness

