## Supplementary material for "Impact of respiratory syncytial virus immunization in older adults: the importance of a valid burden estimation": Online Data Supplement

**Supplementary Table 1**. Case definition of a hospitalization for acute respiratory infection in the hospital-based prospective surveillance network

| Prepandemic seasons  (2012-2013 to 2018-2019) | Admission for ≥24 hours with fever or feverishness in the absence of an identifiable non-respiratory cause, or cough, or sore throat |
| --- | --- |
| Postpandemic seasons  (2020-2021 and beyond) | Admission for ≥24 hours with fever or history of fever not attributed to another illness, or cough [or exacerbation of cough] or difficulty breathing [or exacerbation of difficulty breathing], or sudden extreme fatigue, or at least two of the following symptoms: rhinorrhea or nasal congestion, sore throat, myalgia or arthralgia, or sudden anosmia or ageusia) |

**Supplementary Table 2**. Study eligibility criteria for RSV PCR multiplex validity and final search strategy.

To account for diagnostic test performance in the analysis of HospiVir data, we first reviewed the literature to obtain sensitivity and specificity ranges for RSV testing using multiplex reverse transcriptase polymerase chain reaction (RT-PCR). We searched PubMed MEDLINE from January 2014 through December 2024 for systematic reviews on RSV diagnostic tests accuracy in clinical samples from adults. We then updated the search strategy by including the most recent systematic review of relevant methodological quality^(13)^ using PubMed MEDLINE from January 2022 through December 2024. No additional studies meeting the eligibility criteria were identified in the updated search. Therefore, we retained the sensitivity and specificity estimates reported in the original systematic review with meta-analysis^(13)^ .

|  | **Inclusion criteria** | **Exclusion criteria** |
| --- | --- | --- |
| **Population** | 1. Individuals of all ages with suspected RSV disease from clinical settings | 1. Studies not reporting on patients with RSV disease 2. Studies not reporting results from clinical settings |
| **Exposure** | 3. Diagnostic testing specifications:    3.1. Index test and reference standard available    3.2. Index test is PCR multiplex 4. Specimen specifications:    4.1. RSV specimen types (NP specimen, OP specimen, nasal only specimen, saliva, blood, sputum, and other studied specimen types [alone and in combination, where possible])    4.2. Specimen collection methodology (swab, wash, others)    4.3. Number of specimens tested, per specimen type 5. Laboratory specifications:    5.1. Laboratory storage and collection methods 6. Disease specifications:    6.1. Clinical presentation of RSV    6.2. Time since symptom onset    6.3. Level of care for the RSV disease    6.4. RSV disease severity    6.5. RSV sub-type | 2. Test without reference standard |
| **Outcome** | 6. Diagnostic test performance:    6.1. Sensitivity, specificity, positive predictive value, and negative predictive value for the various levels of exposure    6.2. True positive cases, false positive cases, true negative cases, false negative cases 7. Quantitative comparison of testing methods:    7.1. Number or percentage of patients with RSV infection identified by one RSV testing method versus number or percentage identified by a different testing method in the same population | 3. Studies reporting none of the outcomes of interest |
| **Time (publication periods)** | 8. 2000-2024 | 4. Studies published before 2000 |
| **Study design** | 9. Prospective and retrospective cohort studies, prospective and retrospective cross-sectional studies, case control studies, validation studies, randomized clinical control trials |  |
| **Publication type** | 10. Published articles, online reports, case-series, conference posters or presentations | 5. In vitro studies 6. News and opinion articles 7. Case reports 8. Narrative reviews, letters 9. Systematic literature reviews and meta-analyses |
| **Language** | 11. English, French | 10. Other languages |

Final search strategy adapted from the systematic review by Onwuchekwa et al. (2023), based on selection criteria detailed in Supplementary Material 1: (“Respiratory Tract Infections / diagnosis” OR “sensitivity and specificity”) AND (“Respiratory Syncytial Virus Infections”) OR (“respiratory tract infections / diagnosis” AND “viruses / isolation and purification”) AND (“2000/01/01” : “3000”) AND (“validation study” OR “diagnostic study” OR “diagnostic performance”).

**Supplementary Table 3**. Lash’s equations for bias analysis

**Equation 1**

$$\boldsymbol{B}_{\boldsymbol{1}}\boldsymbol{=(}\boldsymbol{B}_{\boldsymbol{1}}^{\boldsymbol{*}}\boldsymbol{-Fp}\boldsymbol{M}_{\boldsymbol{0}}\boldsymbol{)/(Se+SP-1))}$$

**Equation 2**

$$\boldsymbol{B}_{\boldsymbol{0}}\boldsymbol{=(}\boldsymbol{B}_{\boldsymbol{0}}^{\boldsymbol{*}}\boldsymbol{-Fn}\boldsymbol{M}_{\boldsymbol{0}}\boldsymbol{)/(Se+SP-1))}$$

| **Parameters** | **Definition** |
| --- | --- |
| M_0_ | B_1_*+ B_0_* |
| B_1_* | Number classified as positive |
| B_0_* | Number classified as negative |
| B_1_ | Number truly positive |
| B_0_ | Number truly negative |
| Se | Probability that someone positive is classified as positive = sensitivity = Pr(X* = 1\|X = 1) |
| Fn | Probability that someone positive is classified as negative = false-negative probability = Pr(X* = 0\|X = 1) = 1 − Se |
| Sp | Probability that someone negative is classified as negative = specificity = Pr(X* = 0\|X = 0) |
| Fp | Probability that someone negative is classified as positive = false-positive probability = Pr(X* = 1\|X = 0) = 1 − Sp |

**Supplementary Table 4**. Detailed adjustments to estimate hospitalization incidence in hospital-based prospective surveillance network according to health status and age

| **Season** | **18-64 years** | | **65 years and older** | |
| --- | --- | --- | --- | --- |
|  | **Healthy** | **Comorbidity** | **Healthy** | **Comorbidity** |
| **Adjustment for test validity^a^** | | | | |
| **2012-13** | x0.88 | x0.87 | x0.98 | x0.98 |
| **2013-14** | x0.93 | x0.81 | x0.99 | x0.97 |
| **2014-15** | x0.88 | x0.91 | x0.68 | x0.89 |
| **2015-16** | x0.9 | x0.05 | x0.93 | x0.9 |
| **2016-17** | x1.03 | x1.04 | x1.0 | x1.01 |
| **2017-18** | x0.85 | x0.98 | x0.9 | x1.01 |
| **2018-19** | x0.61 | x0.95 | x1.02 | x1.03 |
| **2022-23** | - | - | x0.67 | x0.67 |
| **2023-24** | - | - | x0.87 | x0.87 |
| **2024-25** | - | - | x0.94 | x0.94 |
| **Extrapolation to the entire season^b^** | | | | |
| **2012-13** | x2.4 | x2.4 | x2.4 | x2.4 |
| **2013-14** | x1.6 | x1.6 | x1.6 | x1.6 |
| **2014-15** | x3.1 | x3.1 | x3.1 | x3.1 |
| **2015-16** | x2.6 | x2.6 | x2.6 | x2.6 |
| **2016-17** | x1.8 | x1.8 | x1.8 | x1.8 |
| **2017-18** | x1.8 | x1.8 | x1.8 | x1.8 |
| **2018-19** | x1.4 | x1.4 | x1.4 | x1.4 |
| **2022-23** | - | - | x1.0 | x1.0 |
| **2023-24** | - | - | x1.0 | x1.0 |
| **2024-25** | - | - | x1.0 | x1.0 |

^a^ Adjustment for sensitivity and specificity of diagnostic test in the Québec prospective hospital-based surveillance network (HospiVir) based on Lash’s equations for bias analysis

^b^ Extrapolation to the entire season according to data from the provincial sentinel laboratory surveillance. Multiplier = 1 / (proportion of positive samples during the weeks included in the surveillance among all positive samples throughout the season)

**Supplementary Table 5**. Exploratory outcomes in health administrative databases (2022-23 to 2024-25 seasons)

| **Exploratory outcomes** | **Definitions** | **Databases** |
| --- | --- | --- |
| RSV-associated emergency department (ED) visits | Visits with an ARI diagnostic code linked to an RSV-positive test 7 days before to 3 days after the visits. | Emergency department database: Système d'information et de gestion des urgences (SIGDU) |
| RSV-associated hospitalization or death after ED visit | Those admitted or deceased after an emergency department visit due to RSV. | SIGDU |
| RSV-associated death (first definition) | Deaths caused by a respiratory infection (ICD-10 codes: J00.x-J22.x, B34.9, or B97.4) and linked to an RSV-positive test within 30 days prior to death. | Deaths registry: Registre des événements démographiques (RED) |
| RSV-associated deaths (second definition) | Deaths caused by RSV infection (ICD-10 codes primary or secondary: J12.1, J20.5, J21.0, or B97.4). | Deaths registry: Registre des événements démographiques (RED) |

Abbreviations: ARI=Acute respiratory infection; ICD-10= International Classification of Disease, Tenth Modification

**Supplementary Table 6**. Adjusted and unadjusted RSV-associated hospitalization rates in hospital-based prospective surveillance network according to age, comorbidity status and season, between 2012-2013 and 2018-2019

|  |  |  | Healthy | | Comorbidity | | Overall | |
| --- | --- | --- | --- | --- | --- | --- | --- | --- |
| Season | Age groups | RSV Positivity (%)* | Adjusted RSV hospitalization rate per 100000 (95% CI) | Rate difference (%) | Adjusted RSV hospitalization rate per 100000 (95% CI) | Rate difference (%) | Adjusted RSV hospitalization rate per 100000 (95% CI) | Rate difference (%) |
| 2012-2013 season |  | | Without adjustment for diagnostic test performance | | | | | |
|  | 18-64 years | 3.1 | NA | NA | 7.6 (0.0 – 16.3) | NA | 2.6 (0.0 – 5.6) | NA |
|  | 65-74 years | 11.1 | NA | NA | 86.9 (1.7 – 172.1) | NA | 39.1 (0.8 – 77.4) | NA |
|  | 75-84 years | 5.2 | 25.0 (0.0 – 74.1) | NA | 111.4 (22.3 – 200.6) | NA | 74.6 (19.3 – 129.9) | NA |
|  | 75+ years | 6.2 | 56.9 (25.0 – 88.7) | NA | 187.2 (138.6 – 235.7) | NA | 133.3 (101.9 – 164.6) | NA |
|  | 85+ years | 7.5 | 156.5 (0.0 – 373.4) | NA | 382.0 (117.3 – 646.7) | NA | 296.5 (112.7 – 480.3) | NA |
|  | Overall | 6.4 | 2.7 (0.0 – 17.2) | NA | 42.9 (20.7 – 65.1) | NA | 15.7 (3.6 – 27.9) | NA |
|  |  | | Adjustment for diagnostic test sensitivity and specificity | | | | | |
|  | 18-64 years | 3.1 | NA | NA | 6.7 (0.0 – 15.1) | -11.8 | 2.3 (0.0 – 3.7) | -11.5 |
|  | 65-74 years | 11.1 | NA | NA | 85.2 (1.7 – 168.6) | -2.0 | 38.3 (23.0 – 53.7) | -2.0 |
|  | 75-84 years | 5.2 | 24.5 (0.0 - 67.7) | -2.0 | 109.2 (21.8 – 196.5) | -2.0 | 73.1 (46.0 – 100.2) | -2.0 |
|  | 75+ years | 6.2 | 55.7 (24.2 – 87.3) | -2.1 | 183.4 (135.3 – 231.5) | -2.0 | 130.6 (99.6 – 161.7) | -2.0 |
|  | 85+ years | 7.5 | 153.4 (0.0 – 344.2) | -2.0 | 374.4 (114.9 – 633.8) | -2.0 | 290.6 (200.5 – 380.7) | -2.0 |
|  | Overall | 6.4 | 2.6 (0.0 – 15.4) | -3.7 | 41.4 (19.6 – 63.2) | -3.5 | 15.2 (12.3 – 18.2) | -3.2 |
| 2013-2014 season |  | | Without adjustment for diagnostic test performance | | | | | |
|  | 18-64 years | 4.0 | 2.2 (0.0 – 4.6) | NA | 5.5 (0.1 – 11.0) | NA | 3.3 (0.9 – 5.8) | NA |
|  | 65-74 years | 5.0 | NA | NA | 30.3 (0.6 – 60.0) | NA | 13.6 (0.3 – 27.0) | NA |
|  | 75-84 years | 9.6 | 13.5 (0.0 – 39.9) | NA | 80.1 (24.6 – 135.6) | NA | 51.7 (17.9 – 85.5) | NA |
|  | 75+ years | 8.6 | 10.1 (0.0 – 23.4) | NA | 85.3 (53.0 – 117.7) | NA | 54.3 (34.5 – 74.0) | NA |
|  | 85+ years | 7.0 | NA | NA | 98.2 (2.0 – 194.4) | NA | 61.0 (1.2 – 120.7) | NA |
|  | Overall | 5.9 | 1.9 (0.0 – 9.6) | NA | 20.1 (9.1 – 31.1) | NA | 7.9 (1.6 – 14.1) | NA |
|  |  | | Adjustment for diagnostic test sensitivity and specificity | | | | | |
|  | 18-64 years | 4.0 | 2.0 (0.0 – 4.3) | -9.1 | 4.8 (0.0 -10.2) | -12.7 | 3.0 (1.4 – 4.5) | -9.1 |
|  | 65-74 years | 5.0 | NA | NA | 29.4 (0.0 – 58.8) | -3.0 | 13.2 (4.4 – 22.1) | -2.9 |
|  | 75-84 years | 9.6 | 13.4 (0.0 – 37.9) | -0.7 | 77.7 (22.7 -132.6) | -3.0 | 50.3 (27.9 – 72.6) | -2.7 |
|  | 75+ years | 8.6 | 10.0 (0.0 – 23.2) | -1.0 | 82.8 (50.9 – 114.7) | -2.9 | 52.7 (33.2 - 72.2) | -2.9 |
|  | 85+ years | 7.0 | NA | NA | 95.2 (0.0 – 190.5) | -3.1 | 59.1 (19.7 – 98.6) | -3.1 |
|  | Overall | 5.9 | 1.8 (0.0 – 8.9) | -5.3 | 19.1 (8.2 – 29.9) | -5.0 | 7.5 (5.4 – 9.5) | -5.1 |
| 2014-2015 season |  | | Without adjustment for diagnostic test performance | | | | | |
|  | 18-64 years | 3.8 | NA | NA | 10.8 (0.2 -21.4) | NA | 3.7 (0.1 – 7.3) | NA |
|  | 65-74 years | 6.0 | NA | NA | 113.9 (35.0 – 192.9) | NA | 51.3 (15.7 – 86.8) | NA |
|  | 75-84 years | 5.1 | 18.3 (0.0 – 68.8) | NA | 191.5 (72.8 – 310.2) | NA | 117.7 (46.2 – 189.2) | NA |
|  | 75+ years | 3.8 | 13.5 (0.0 – 28.6) | NA | 187.9 (140.7 – 235.2) | NA | 116.0 (87.5 – 144.4) | NA |
|  | 85+ years | 2.2 | NA | NA | 179.6 (3.6 – 355.5) | NA | 111.5 (2.2 – 220.8) | NA |
|  | Overall | 4.2 | 0.7 (0.0 – 14.3) | NA | 50.4 (30.0 -70.9) | NA | 17.0 (5.6 – 28.4) | NA |
|  |  | | Adjustment for diagnostic test sensitivity and specificity | | | | | |
|  | 18-64 years | 3.8 | NA | NA | 9.8 (2.6 – 17.0) | -9.3 | 3.4 (1.7 – 5.0) | -8.2 |
|  | 65-74 years | 6.0 | NA | NA | 101.4 (47.7 – 155.1) | -11.0 | 45.6 (29.5 – 61.7) | -11.1 |
|  | 75-84 years | 5.1 | 18.3 (0.0 – 62.8) | 0.0 | 170.5 (89.7 – 251.2) | -11.0 | 105.6 (73.6 – 137.7) | -10.3 |
|  | 75+ years | 3.8 | 13.5 (0.0 – 28.6) | 0.0 | 167.3 (122.7 – 211.9) | -11.0 | 103.8 (76.9 – 130.7) | -10.5 |
|  | 85+ years | 2.2 | NA | NA | 159.8 (40.2 – 279.5) | -11.0 | 99.2 (49.8 – 148.7) | -11.0 |
|  | Overall | 4.2 | 0.7 (0.0 – 12.7) | 0.0 | 45.1 (31.1 – 59.0) | -10.5 | 15.2 (12.3 – 18.2) | -10.6 |
| 2015-2016 season |  | | Without adjustment for diagnostic test performance | | | | | |
|  | 18-64 years | 1.7 | 4.0 (0.0 – 8.6) | NA | NA | NA | 2.6 (0.0 – 5.6) | NA |
|  | 65-74 years | 5.4 | 10.7 (0.0 – 31.8) | NA | 65.6 (8.1 – 123.0) | NA | 35.4 (7.1 – 63.7) | NA |
|  | 75-84 years | 6.9 | 24.1 (0.0 -71.4) | NA | 161.2 (55.9 -266.5) | NA | 102.8 (39.1 – 166.5) | NA |
|  | 75+ years | 4.2 | NA | NA | 111.8 (75.9 – 147.8) | NA | 73.0 (50.8 – 95.3) | NA |
|  | 85+ years | 0.0 | NA | NA | NA | NA | NA | NA |
|  | Overall | 3.6 | 3.6 (0.0 – 15.6) | NA | 25.6 (10.4 – 40.9) | NA | 10.9 (1.4 – 20.4) | NA |
|  |  | | Adjustment for diagnostic test sensitivity and specificity | | | | | |
|  | 18-64 years | 1.7 | 3.6 (0.0 – 7.7) | -10.0 | NA | NA | 2.4 (1.0 -3.8) | -7.7 |
|  | 65-74 years | 5.4 | 10.0 (0.0 – 28.9) | -6.5 | 59.0 (5.6 – 112.5) | -10.1 | 32.0 (18.8 – 45.3) | -9.6 |
|  | 75-84 years | 6.9 | 22.4 (0.0 – 65.0) | -7.1 | 145.1 (47.1 – 243.0) | -10.0 | 92.8 (63.0 – 122.6) | -9.7 |
|  | 75+ years | 4.2 | NA | NA | NA | NA | NA | NA |
|  | 85+ years | 0.0 | NA | NA | NA | NA | NA | NA |
|  | Overall | 3.6 | 4.1 (0.0 – 14.9) |  | 23.1 (8.9 – 37.2) | -9.8 | 10.4 (7.9 – 12.8) | -4.6 |
| 2016-2017 season |  | | Without adjustment for diagnostic test performance | | | | | |
|  | 18-64 years | 15.2 | 3.8 (0.5 – 7.2) | NA | 26.3 (14.2 – 38.5) | NA | 11.5 (6.8 – 16.2) | NA |
|  | 65-74 years | 15.7 | 11.2 (0.0 – 26.7) | NA | 123.2 (66.3 – 180.1) | NA | 61.6 (34.6 – 88.6) | NA |
|  | 75-84 years | 7.4 | NA | NA | 119.0 (54.3 – 183.7) | NA | 68.3 (31.2 – 105.4) | NA |
|  | 75+ years | 9.5 | 26.6 (6.3 – 47.0) | NA | 192.6 (146.8 – 238.4) | NA | 124.3 (96.1 – 152.5) | NA |
|  | 85+ years | 11.5 | 95.0 (0.0 – 202.6) | NA | 348.1 (187.3 – 508.8) | NA | 252.2 (144.3 – 360.0) | NA |
|  | Overall | 12.1 | 4.2 (0.0 – 14.7) | NA | 67.6 (50.9 – 84.3) | NA | 25.7 (16.8 – 34.7) | NA |
|  |  | | Adjustment for diagnostic test sensitivity and specificity | | | | | |
|  | 18-64 years | 15.2 | 3.9 (0.5 – 7.4) | 2.6 | 27.4 (15.2 – 39.5) | 4.2 | 12.0 (8.7 – 15.2) | 4.3 |
|  | 65-74 years | 15.7 | 11.2 (0.0 – 27.2) | 0.0 | 124.4 (67.5 – 181.3) | 1.0 | 62.1 (43.8 – 80.4) | 0.8 |
|  | 75-84 years | 7.4 | NA | NA | 120.2 (55.5 -184.9) | 1.0 | 69.0 (43.8 – 94.2) | 1.0 |
|  | 75+ years | 9.5 | 26.6 (6.3 – 47.0) | 0.0 | 194.5 (148.5 – 240.5) | 1.0 | 125.4 (97.1 – 153.7) | 0.9 |
|  | 85+ years | 11.5 | 95.0 (0.0 – 205.8) | 0.0 | 351.5 (190.7 – 512.3) | 1.0 | 254.3 (181.2 -327.4) | 0.8 |
|  | Overall | 12.1 | 5.3 (0.0 – 16.1) | 26.2 | 68.8 (52.2 – 85.5) | 1.8 | 26.9 (22.9 – 30.9) | 4.7 |
| 2017-2018 season |  | | Without adjustment for diagnostic test performance | | | | | |
|  | 18-64 years | 6.0 | NA | NA | 11.8 (3.6 – 19.9) | NA | 4.0 (1.2 – 6.8) | NA |
|  | 65-74 years | 7.2 | NA | NA | 66.7 (25.4 – 108.0) | NA | 30.0 (11.4 – 48.6) | NA |
|  | 75-84 years | 11.7 | 24.4 (0.0 – 58.3) | NA | 172.2 (94.8 – 249.6) | NA | 109.2 (62.5 – 156.0) | NA |
|  | 75+ years | 11.0 | 17.4 (1.2 – 33.7) | NA | 219.3 (171.2 – 267.5) | NA | 136.3 (107.1 – 165.4) | NA |
|  | 85+ years | 10.3 | NA | NA | 315.9 (165.7 – 466.0) | NA | 196.2 (102.9 – 289.4) | NA |
|  | Overall | 9.1 | 1.0 (0.0 – 10.0) | NA | 54.4 (38.7 – 70.0) | NA | 19.2 (11.3 – 27.2) | NA |
|  |  | | Adjustment for diagnostic test sensitivity and specificity | | | | | |
|  | 18-64 years | 6.0 | NA | NA | 11.5 (4.2 – 18.9) | -2.5 | 4.0 (2.1 – 5.8) | 0.0 |
|  | 65-74 years | 7.2 | NA | NA | 67.4 (30.2 – 104.6) | 1.0 | 30.3 (17.7 – 42.9) | 1.0 |
|  | 75-84 years | 11.7 | 22.0 (0.0 – 50.8) | -9.8 | 173.9 (104.2 – 243.6) | 1.0 | 109.2 (77.8 – 140.6) | 0.0 |
|  | 75+ years | 11.0 | NA | NA | 221.5 (173.1 -269.9) | 1.0 | 136.8 (107.7 – 166.0) | 0.4 |
|  | 85+ years | 10.3 | NA | NA | 319.0 (183.9 – 454.2) | 1.0 | 198.1 (135.1 – 261.1) | 1.0 |
|  | Overall | 9.1 | 0.9 (0.0 – 8.6) | -10.0 | 54.7 (40.6 – 68.8) | 0.6 | 19.3 (15.9 – 22.7) | 0.5 |
| 2018-2019 season |  | | Without adjustment for diagnostic test performance | | | | | |
|  | 18-64 years | 9.9 | 0.8 (0.0 – 2.2) | NA | 11.6 (3.6 – 19.7) | NA | 4.5 (1.6 – 7.4) | NA |
|  | 65-74 years | 6.8 | NA | NA | 91.5 (48.0 – 135.1) | NA | 41.2 (21.6 – 60.8) | NA |
|  | 75-84 years | 10.5 | 35.1 (0.0 – 74.8) | NA | 156.3 (84.1 – 228.5) | NA | 104.7 (59.9 – 149.4) | NA |
|  | 75+ years | 10.5 | 50.5 (18.7 – 82.4) | NA | 218.0 (162.6 – 273.3) | NA | 149.0 (113.9 -184.1) | NA |
|  | 85+ years | 10.5 | 90.1 (0.0 – 192.0) | NA | 348.2 (191.6 – 504.8) | NA | 250.4 (145.8 – 355.0) | NA |
|  | Overall | 11.4 | 3.5 (0.0 – 13.9) | NA | 59.4 (42.9 – 75.9) | NA | 22.9 (14.0 – 31.7) | NA |
|  |  | | Adjustment for diagnostic test sensitivity and specificity | | | | | |
|  | 18-64 years | 9.9 | 0.5 (0.0 – 1.4) | -37.5 | 11.0 (2.8 – 19.3) | -5.2 | 4.1 (1.9 – 6.3) | -8.9 |
|  | 65-74 years | 6.8 | NA | NA | 94.3 (49.9 – 138.7) | 3.1 | 42.4 (26.8 – 58.1) | 2.9 |
|  | 75-84 years | 10.5 | 35.8 (11.6 – 60.0) | 2.0 | 161.0 (87.3 – 234.6) | 3.0 | 107.6 (71.9 – 143.4) | 2.8 |
|  | 75+ years | 10.5 | 51.5 (19.4 – 83.7) | 2.0 | 224.5 (168.3 – 280.7) | 3.0 | 153.3 (117.7 – 188.9) | 2.9 |
|  | 85+ years | 10.5 | 91.9 (29.7 – 154.1) | 2.0 | 358.7 (199.0 – 518.4) | 3.0 | 257.6 (173.9 -341.2) | 2.9 |
|  | Overall | 11.4 | 3.4 (0.0 – 9.7) | -2.9 | 60.6 (43.8 – 77.4) | 2.0 | 23.2 (18.9 – 27.5) | 1.3 |

NA: not applicable; CI : confidence interval; RSV; respiratory syncytial virus

**Supplementary Table 7**. Observed and expected RSV-associated hospitalizations in hospital-based prospective surveillance network following the RSV immunization campaign in adults aged 75 years and older in Québec, Canada, using 2 pasts seasons (2022-2023 and 2023-2024) as reference

| **Burden estimation method** | **Targeted group** | **Observed RSV-associated hospitalizations, n** | **Expected RSV-associated hospitalizations, n (95% CI)** | **Absolute impact measure, n (95 %CI)** | **Relative impact measure, % (95 %CI)** | **P value**  **(Exact Poisson)** | **Impact (Standardized Morbidity Ratio), % (95 %CI)a** |
| --- | --- | --- | --- | --- | --- | --- | --- |
|  | Group used to estimate the difference or ratio for the targeted group**: individuals aged 65-74 years  N_2023-24_ = 26; N_2024-25_ = 34 | | | | | |  |
| Without adjustment for diagnostic test performance | All 75+ years | 66 | 94 (56 ; 167) | -28 (-101 ; 10) | -30 (-60 ; 17) | 0.003 | -30 (-46 ; -11) |
|  | All 80+ years | 49 | 71 (44 ; 124) | -22 (-75 ; 5) | -31 (-61 ; 12) | 0.009 | -31 (-49 ; -8) |
|  | All 85+ years | 30 | 44 (33 ; 66) | -14 (-36 ; -3) | -33 (-55 ; -10) | 0.03 | -33 (-54 ; -4) |
| Adjustment for diagnostic test sensitivity and specificity | All 75+ years | 62 | 87 (52 ; 157) | -25 (-95 ; 10) | -29 (-60 ; 20) | 0.004 | -30 (-46 ; -10) |
|  | All 80+ years | 46 | 66 (40 ; 120) | -20 (-74 ; 6) | -30 (-61 ; 14) | 0.009 | -31 (-50 ; -8) |
|  | All 85+ years | 28 | 41 (23 ; 78) | -13 (-50 ; 5) | -33 (-64 ; -21) | 0.04 | -32 (-55 ; -2) |

PSR: private senior homes; CI: confidence interval

**ajouter méthode

**^a^SMR IC 95% exact confidence interval (Clopper-Pearson)** based on Poisson regression

**Supplementary Table 8.** Changes in incidence of RSV-associated hospitalisations among older adults based on health administrative data in Québec, Canada (2022-2023 to 2024-2025) by age group

| Outcome | **Rate† or proportion in 65 to 74**  **years old (95% CI)** | | | **Rate† or proportion in ≥75**  **years old (95% CI)** | | | **Rate† or proportion in ≥80**  **years old with comorbidities (95% CI)** | | | **Rate† or proportion in ≥85**  **years old with comorbidities (95% CI)** | | |
| --- | --- | --- | --- | --- | --- | --- | --- | --- | --- | --- | --- | --- |
|  | **Pre-intervention** | **Post-intervention** | **Post/pre ratio** | **Pre-intervention** | **Post-intervention** | **Post/pre ratio** | **Pre-intervention** | **Post-intervention** | **Post/pre ratio** | **Pre-intervention** | **Post-intervention** | **Post/pre ratio** |
| RSV-associated ED visit (rate per 100 000 person-years) | 34.0  (31.5 - 36.5) | 75.1  (69.9 - 80.4) | 2.21 (2.06 - 2.37) | 92.2  (87.6 - 96.8) | 182.8  (173.7 - 191.9) | 1.98  (1.89 - 2.09) | 131.3  (123.5 - 139.2) | 245.2  (230.2 - 260.2) | 1.87  (1.76 - 1.99) | 168.6  (156.2 - 180.9) | 278.0  (255.6 - 300.4) | 1.65  (1.52 - 1.79) |
| RSV-associated hospitalization or death after ED visit (%) | 46.2  (42.5 - 49.9) | 50.1  (46.6 - 53.5) | 1.08 (0.98 - 1.21) | 54.6  (52.1 - 57.1) | 56.8  (54.3 - 59.2) | 1.04  (0.98 - 1.11) | 57.5  (54.6 - 60.4) | 58.9  (55.9 - 61.9) | 1.02  (0.95 - 1.10) | 59.0  (55.4 - 62.6) | 62.7  (58.8 - 66.5) | 1.06  (0.97 - 1.16) |
| RSV-associated death (first definition) (rate per 100 000 person-years) | 1.8  (1.2 - 2.4) | 3.0  (2.0 - 4.1) | 1.67 (1.18 - 2.36) | 17.2  (15.2 - 19.1) | 24.4  (21.1 - 27.7) | 1.42  (1.24 - 1.63) | 29.8  (26.1 - 33.5) | 38.4  (32.5 - 44.4) | 1.29  (1.11 - 1.51) | 44.6  (38.2 - 50.9) | 54.4  (44.5 - 64.3) | 1.22  (1.02 - 1.46) |
| RSV-associated deaths (second definition) (rate per 100 000 person-years) | 1.3  (0.8 - 1.7) | 2.4  (1.4 - 3.3) | 1.90 (1.29 - 2.81) | 14.2  (12.4 - 16.0) | 14.9  (12.3 - 17.5) | 1.05  (0.88 - 1.25) | 24.9  (21.5 - 28.3) | 22.9  (18.3 - 27.5) | 0.92  (0.75 - 1.12) | 35.8  (30.1 - 41.5) | 35.2  (27.2 - 43.1) | 0.98  (0.78 - 1.23) |

CI: confidence interval; RSV: Respiratory syncytial virus.

**Supplementary Table 9.** By age group, changes in observed number of RSV associated outcomes among older adults compared to the expected number in counterfactual situation without the immunization campaign, based on health administrative data in Québec, Canada (2022-2023 to 2024-2025)

| **Outcome** | | **Reduction adjusted for time trends in younger adults (%, 95% CI)** | **Rates or proportions post-intervention** | | **Number of severe cases post-intervention** | | **Number of prevented cases** |
| --- | --- | --- | --- | --- | --- | --- | --- |
|  |  |  | **Observed** | **Expected**  **(95% CI)** | **Observed** | **Expected**  **(95% CI)** |  |
| ≥75 years old | RSV-associated ED visit (rate per 100 000 person-years) | 10%  (-2% – 21%) | 182.81 | 203.67  (179.98 – 230.44) | 1558 | 1736  (1534 – 1964) | 178 ED visits  (-24 – 406)  105 admissions  (-14 – 240) |
|  | RSV-associated hospitalization or death after ED visit (%) | 4%  (-10% – 17%) | 0.57 | 0.59  (0.51 – 0.68) | 885 | 923  (802 – 1063) | 38 admissions  (-83 – 178) |
|  | RSV-associated death (first definition) (rate per 100 000 person-years) | 45%  (1% – 69%) | 14.90 | 27.06  (15.01 – 48.81) | 127 | 231  (128 – 416) | 104 deaths  (1 – 289) |
|  | RSV-associated deaths (second definition) (rate per 100 000 person-years) | 15%  (-41% – 48%) | 24.41 | 28.58  (17.28 – 47.26) | 208 | 244  (147 – 403) | 36 deaths  (-61 – 195) |
| ≥80 years old with comorbidities | RSV-associated ED visit (rate per 100 000 person-years) | 16%  (4% – 26%) | 245.24 | 290.26  (254.21 – 331.40) | 1027 | 1216  (1065 – 1388) | 189 ED visits  (38 – 361)  117 admissions  (24 – 224) |
|  | RSV-associated hospitalization or death after ED visit (%) | 5%  (-9% – 18%) | 0.59 | 0.62  (0.54 – 0.72) | 605 | 639  (555 – 736) | 34 admissions  (-50 – 131) |
|  | RSV-associated death (first definition) (rate per 100 000 person-years) | 52%  (12% – 73%) | 22.92 | 47.41  (26.02 – 86.41) | 96 | 199  (109 – 362) | 103 deaths  (13 – 266) |
|  | RSV-associated deaths (second definition) (rate per 100 000 person-years) | 22%  (-29% – 53%) | 38.45 | 49.59  (29.76 – 82.63) | 161 | 208  (125 – 346) | 47 deaths  (-36 – 185) |
| ≥85 years old with comorbidities | RSV-associated ED visit (rate per 100 000 person-years) | 25%  (13% – 36%) | 277.99 | 372.49  (320.97 – 432.27) | 593 | 795  (685 – 922) | 202 ED visits  (92 – 329)  129 admissions  (59 – 211) |
|  | RSV-associated hospitalization or death after ED visit (%) | 3%  (-14% – 17%) | 0.63 | 0.64  (0.55 – 0.75) | 372 | 382  (326 – 447) | 10 admissions  (-46 – 75) |
|  | RSV-associated death (first definition) (rate per 100 000 person-years) | 48%  (5% – 72%) | 35.16 | 68.14  (36.84 – 125.97) | 75 | 145  (79 – 269) | 70 deaths  (4 – 194) |
|  | RSV-associated deaths (second definition) (rate per 100 000 person-years) | 27%  (-24% – 57%) | 54.38 | 74.20  (43.93 – 125.30) | 116 | 158  (94 – 267) | 42 deaths  (-22 – 151) |

CI: confidence interval; RSV: Respiratory syncytial virus;

**Supplementary Table 10**. Number needed to vaccinate and avertable hospitalizations according to health status and age

| Season | Age groups | RSV hospitalization rate per 100000 | Vaccine effectiveness (%) | Number needed vaccinate | Avertable hospitalizations | | | |
| --- | --- | --- | --- | --- | --- | --- | --- | --- |
|  | All Patients |  |  |  | VC=10% | VC=30% | VC=60% | VC=90% |
| 2012-2013 season | 18-64 y | 2.28 | 70 | 62,617 | 8 | 25 | 50 | 75 |
|  |  |  | 80 | 54,789 | 10 | 29 | 57 | 86 |
|  |  |  | 90 | 48,701 | 11 | 32 | 65 | 97 |
|  | 65-74 y | 38.32 | 70 | 3,728 | 20 | 59 | 117 | 176 |
|  |  |  | 80 | 3,262 | 22 | 67 | 134 | 201 |
|  |  |  | 90 | 2,899 | 25 | 76 | 151 | 227 |
|  | 75-84 y | 73.11 | 70 | 1,954 | 21 | 63 | 126 | 189 |
|  |  |  | 80 | 1,710 | 24 | 72 | 144 | 216 |
|  |  |  | 90 | 1,520 | 27 | 81 | 162 | 243 |
|  | 75+y | 130.62 | 70 | 1,094 | 54 | 162 | 323 | 485 |
|  |  |  | 80 | 957 | 62 | 185 | 369 | 554 |
|  |  |  | 90 | 851 | 69 | 208 | 415 | 623 |
|  | 85+ y | 290.60 | 70 | 492 | 33 | 99 | 197 | 296 |
|  |  |  | 80 | 430 | 38 | 113 | 225 | 338 |
|  |  |  | 90 | 382 | 42 | 127 | 253 | 380 |
|  | Patients with comorbidity |  |  |  |  |  |  |  |
|  | 18-64 y | 6.65 | 70 | 21,477 | 8 | 25 | 50 | 75 |
|  |  |  | 80 | 18,793 | 10 | 29 | 57 | 86 |
|  |  |  | 90 | 16,075 | 11 | 32 | 65 | 97 |
|  | 65-74 y | 85.16 | 70 | 1,678 | 20 | 59 | 117 | 176 |
|  |  |  | 80 | 1,468 | 22 | 67 | 134 | 201 |
|  |  |  | 90 | 1,305 | 25 | 76 | 151 | 227 |
|  | 75-84 y | 109.18 | 70 | 1,308 | 18 | 54 | 108 | 162 |
|  |  |  | 80 | 1,145 | 21 | 62 | 123 | 185 |
|  |  |  | 90 | 1,018 | 23 | 69 | 139 | 208 |
|  | 75+ y | 183.43 | 70 | 779 | 44 | 133 | 266 | 398 |
|  |  |  | 80 | 681 | 51 | 152 | 304 | 455 |
|  |  |  | 90 | 606 | 57 | 171 | 342 | 512 |
|  | 85+ y | 374.37 | 70 | 382 | 26 | 79 | 158 | 236 |
|  |  |  | 80 | 334 | 30 | 90 | 180 | 270 |
|  |  |  | 90 | 297 | 34 | 101 | 203 | 304 |
|  | All Patients |  |  |  |  |  |  |  |
| 2013-2014 season | 18-64 y | 2.98 | 70 | 48,003 | 11 | 33 | 66 | 98 |
|  |  |  | 80 | 42,003 | 12 | 37 | 75 | 112 |
|  |  |  | 90 | 37,336 | 14 | 42 | 84 | 126 |
|  | 65-74 y | 13.23 | 70 | 10,800 | 7 | 21 | 42 | 64 |
|  |  |  | 80 | 9,450 | 8 | 24 | 48 | 73 |
|  |  |  | 90 | 8,400 | 9 | 27 | 54 | 82 |
|  | 75-84 y | 50.29 | 70 | 2,841 | 15 | 44 | 87 | 131 |
|  |  |  | 80 | 2,486 | 17 | 50 | 100 | 150 |
|  |  |  | 90 | 2,210 | 19 | 56 | 112 | 169 |
|  | 75+ y | 52.71 | 70 | 2,710 | 21 | 64 | 129 | 193 |
|  |  |  | 80 | 2,372 | 25 | 74 | 147 | 221 |
|  |  |  | 90 | 2,108 | 28 | 83 | 166 | 249 |
|  | 85+ y | 59.13 | 70 | 2,416 | 7 | 21 | 42 | 63 |
|  |  |  | 80 | 2,114 | 8 | 24 | 48 | 72 |
|  |  |  | 90 | 1,879 | 9 | 27 | 54 | 81 |
|  | Patients with comorbidity |  |  |  |  |  |  |  |
|  | 18-64 y | 4.82 | 70 | 29,666 | 6 | 18 | 36 | 55 |
|  |  |  | 80 | 25,957 | 7 | 21 | 42 | 62 |
|  |  |  | 90 | 23,073 | 8 | 23 | 47 | 70 |
|  | 65-74 y | 29.40 | 70 | 4,860 | 7 | 21 | 42 | 64 |
|  |  |  | 80 | 4,252 | 8 | 24 | 48 | 73 |
|  |  |  | 90 | 3,780 | 9 | 27 | 54 | 82 |
|  | 75-84 y | 77.70 | 70 | 1,839 | 13 | 39 | 78 | 116 |
|  |  |  | 80 | 1,609 | 15 | 44 | 89 | 133 |
|  |  |  | 90 | 1,430 | 17 | 50 | 100 | 150 |
|  | 75+ y | 82.77 | 70 | 1,726 | 20 | 60 | 119 | 179 |
|  |  |  | 80 | 1,510 | 23 | 68 | 136 | 205 |
|  |  |  | 90 | 1,342 | 26 | 77 | 153 | 230 |
|  | 85+ y | 95.22 | 70 | 1,500 | 7 | 21 | 42 | 63 |
|  |  |  | 80 | 1,313 | 8 | 24 | 48 | 72 |
|  |  |  | 90 | 1,167 | 9 | 27 | 54 | 81 |
|  | All Patients |  |  |  |  |  |  |  |
| 2014-2015 season | 18-64 y | 3.37 | 70 | 42,416 | 12 | 37 | 74 | 111 |
|  |  |  | 80 | 37,114 | 14 | 42 | 85 | 127 |
|  |  |  | 90 | 32,991 | 16 | 48 | 95 | 143 |
|  | 65-74 y | 45.63 | 70 | 3,131 | 25 | 76 | 152 | 228 |
|  |  |  | 80 | 2,739 | 29 | 87 | 174 | 261 |
|  |  |  | 90 | 2,435 | 33 | 98 | 196 | 293 |
|  | 75-84 y | 105.61 | 70 | 1,353 | 31 | 93 | 187 | 280 |
|  |  |  | 80 | 1,184 | 36 | 107 | 213 | 320 |
|  |  |  | 90 | 1,052 | 40 | 120 | 240 | 360 |
|  | 75+ y | 103.81 | 70 | 1,376 | 43 | 130 | 260 | 389 |
|  |  |  | 80 | 1,204 | 49 | 148 | 297 | 445 |
|  |  |  | 90 | 1,070 | 56 | 167 | 334 | 501 |
|  | 85+ y | 99.24 | 70 | 1,439 | 12 | 36 | 73 | 109 |
|  |  |  | 80 | 1,260 | 14 | 42 | 83 | 125 |
|  |  |  | 90 | 1,120 | 16 | 47 | 94 | 140 |
|  | Patients with comorbidity |  |  |  |  |  |  |  |
|  | 18-64 y | 9.82 | 70 | 14,549 | 12 | 37 | 74 | 111 |
|  |  |  | 80 | 12,730 | 14 | 42 | 85 | 127 |
|  |  |  | 90 | 11,316 | 16 | 48 | 95 | 143 |
|  | 65-74 y | 101.40 | 70 | 1,409 | 25 | 76 | 152 | 228 |
|  |  |  | 80 | 1,233 | 29 | 87 | 174 | 261 |
|  |  |  | 90 | 1,096 | 33 | 98 | 196 | 293 |
|  | 75-84 y | 170.45 | 70 | 838 | 29 | 86 | 173 | 259 |
|  |  |  | 80 | 733 | 33 | 99 | 198 | 296 |
|  |  |  | 90 | 652 | 37 | 111 | 222 | 333 |
|  | 75+ y | 167.27 | 70 | 854 | 41 | 123 | 246 | 368 |
|  |  |  | 80 | 747 | 47 | 140 | 281 | 421 |
|  |  |  | 90 | 664 | 53 | 158 | 316 | 474 |
|  | 85+ y | 159.81 | 70 | 894 | 12 | 36 | 73 | 109 |
|  |  |  | 80 | 782 | 14 | 42 | 83 | 125 |
|  |  |  | 90 | 695 | 16 | 47 | 94 | 140 |
|  | All Patients |  |  |  |  |  |  |  |
| 2015-2016 season | 18-64 y | 2.38 | 70 | 60,066 | 9 | 26 | 52 | 78 |
|  |  |  | 80 | 52,557 | 10 | 30 | 60 | 89 |
|  |  |  | 90 | 46,718 | 11 | 34 | 67 | 101 |
|  | 65-74 y | 32.04 | 70 | 4,458 | 18 | 55 | 111 | 166 |
|  |  |  | 80 | 3,901 | 21 | 63 | 127 | 190 |
|  |  |  | 90 | 3,468 | 24 | 71 | 142 | 214 |
|  | 75-84 y | 92.83 | 70 | 1,539 | 28 | 83 | 166 | 249 |
|  |  |  | 80 | 1,347 | 32 | 95 | 190 | 285 |
|  |  |  | 90 | 1,197 | 36 | 107 | 214 | 320 |
|  | 75+ y | 65.94 | 70 | 2,167 | 28 | 84 | 168 | 252 |
|  |  |  | 80 | 1,896 | 32 | 96 | 192 | 288 |
|  |  |  | 90 | 1,685 | 36 | 108 | 216 | 324 |
|  | 85+ y | NA | 70 | NA | NA | NA | NA | NA |
|  |  |  | 80 | NA | NA | NA | NA | NA |
|  |  |  | 90 | NA | NA | NA | NA | NA |
|  | Patients with comorbidity |  |  |  |  |  |  |  |
|  | 18-64 y | NA | 70 | NA | NA | NA | NA | NA |
|  |  |  | 80 | NA | NA | NA | NA | NA |
|  |  |  | 90 | NA | NA | NA | NA | NA |
|  | 65-74 y | 59.01 | 70 | 2,421 | 15 | 46 | 92 | 138 |
|  |  |  | 80 | 2,118 | 17 | 52 | 105 | 157 |
|  |  |  | 90 | 1,883 | 20 | 59 | 118 | 177 |
|  | 75-84 y | 145.06 | 70 | 985 | 25 | 75 | 149 | 224 |
|  |  |  | 80 | 862 | 28 | 85 | 170 | 256 |
|  |  |  | 90 | 766 | 32 | 96 | 192 | 287 |
|  | 75+ y | 100.66 | 70 | 1,419 | NA | NA | NA | NA |
|  |  |  | 80 | 1,242 | NA | NA | NA | NA |
|  |  |  | 90 | 1,104 | NA | NA | NA | NA |
|  | 85+ y | NA | 70 | NA | NA | NA | NA | NA |
|  |  |  | 80 | NA | NA | NA | NA | NA |
|  |  |  | 90 | NA | NA | NA | NA | NA |
|  | All Patients |  |  |  |  |  |  |  |
| 2016-2017 season | 18-64 y | 11.97 | 70 | 11,933 | 44 | 131 | 262 | 393 |
|  |  |  | 80 | 10,441 | 50 | 150 | 299 | 449 |
|  |  |  | 90 | 9,281 | 56 | 168 | 337 | 505 |
|  | 65-74 y | 62.13 | 70 | 2,299 | 37 | 111 | 222 | 334 |
|  |  |  | 80 | 2,012 | 42 | 127 | 254 | 381 |
|  |  |  | 90 | 1,788 | 48 | 143 | 286 | 429 |
|  | 75-84 y | 68.99 | 70 | 2,071 | 21 | 63 | 126 | 190 |
|  |  |  | 80 | 1,812 | 24 | 72 | 145 | 217 |
|  |  |  | 90 | 1,611 | 27 | 81 | 163 | 244 |
|  | 75+ y | 125.41 | 70 | 1,139 | 55 | 165 | 329 | 494 |
|  |  |  | 80 | 997 | 63 | 188 | 376 | 564 |
|  |  |  | 90 | 886 | 71 | 212 | 423 | 635 |
|  | 85+ y | 254.33 | 70 | 562 | 34 | 101 | 201 | 302 |
|  |  |  | 80 | 491 | 38 | 115 | 230 | 345 |
|  |  |  | 90 | 437 | 43 | 129 | 259 | 388 |
|  | Patients with comorbidity |  |  |  |  |  |  |  |
|  | 18-64 y | 27.37 | 70 | 5,219 | 34 | 103 | 205 | 308 |
|  |  |  | 80 | 4,567 | 39 | 117 | 235 | 352 |
|  |  |  | 90 | 4,059 | 44 | 132 | 264 | 396 |
|  | 65-74 y | 124.39 | 70 | 1,148 | 33 | 100 | 200 | 301 |
|  |  |  | 80 | 1,005 | 38 | 114 | 229 | 343 |
|  |  |  | 90 | 893 | 43 | 129 | 258 | 386 |
|  | 75-84 y | 120.19 | 70 | 1,189 | 21 | 63 | 126 | 190 |
|  |  |  | 80 | 1,040 | 24 | 72 | 145 | 217 |
|  |  |  | 90 | 924 | 27 | 81 | 163 | 244 |
|  | 75+ y | 194.52 | 70 | 734 | 50 | 150 | 299 | 449 |
|  |  |  | 80 | 643 | 57 | 171 | 342 | 513 |
|  |  |  | 90 | 571 | 64 | 192 | 385 | 577 |
|  | 85+ y | 351.53 | 70 | 406 | 29 | 86 | 173 | 259 |
|  |  |  | 80 | 356 | 33 | 99 | 197 | 296 |
|  |  |  | 90 | 316 | 37 | 111 | 222 | 333 |
|  | All Patients |  |  |  |  |  |  |  |
| 2017-2018 season | 18-64 y | 3.96 | 70 | 36,082 | 14 | 43 | 87 | 130 |
|  |  |  | 80 | 31,571 | 17 | 50 | 99 | 149 |
|  |  |  | 90 | 28,063 | 19 | 56 | 112 | 167 |
|  | 65-74 y | 30.31 | 70 | 4,713 | 19 | 56 | 112 | 168 |
|  |  |  | 80 | 4,124 | 21 | 64 | 128 | 192 |
|  |  |  | 90 | 3,666 | 24 | 72 | 144 | 216 |
|  | 75-84 y | 109.20 | 70 | 1,308 | 34 | 103 | 207 | 310 |
|  |  |  | 80 | 1,145 | 39 | 118 | 236 | 354 |
|  |  |  | 90 | 1,018 | 44 | 133 | 266 | 398 |
|  | 75+ y | 136.83 | 70 | 1,044 | 62 | 186 | 371 | 557 |
|  |  |  | 80 | 914 | 71 | 212 | 424 | 637 |
|  |  |  | 90 | 812 | 80 | 239 | 478 | 716 |
|  | 85+ y | 198.13 | 70 | 721 | 27 | 82 | 163 | 245 |
|  |  |  | 80 | 631 | 31 | 93 | 186 | 280 |
|  |  |  | 90 | 561 | 35 | 105 | 210 | 315 |
|  | Patients with comorbidity |  |  |  |  |  |  |  |
|  | 18-64 y | 11.54 | 70 | 12,376 | 14 | 43 | 87 | 130 |
|  |  |  | 80 | 10,829 | 17 | 50 | 99 | 149 |
|  |  |  | 90 | 9,626 | 19 | 56 | 112 | 167 |
|  | 65-74 y | 67.36 | 70 | 2,121 | 19 | 56 | 112 | 168 |
|  |  |  | 80 | 1,856 | 21 | 64 | 128 | 192 |
|  |  |  | 90 | 1,649 | 24 | 72 | 144 | 216 |
|  | 75-84 y | 173.92 | 70 | 821 | 31 | 94 | 189 | 283 |
|  |  |  | 80 | 719 | 36 | 108 | 216 | 324 |
|  |  |  | 90 | 639 | 40 | 121 | 243 | 364 |
|  | 75+ y | 221.50 | 70 | 645 | 59 | 176 | 352 | 528 |
|  |  |  | 80 | 564 | 67 | 201 | 402 | 603 |
|  |  |  | 90 | 502 | 75 | 226 | 452 | 679 |
|  | 85+ y | 319.04 | 70 | 448 | 27 | 82 | 163 | 245 |
|  |  |  | 80 | 392 | 31 | 93 | 186 | 280 |
|  |  |  | 90 | 348 | 35 | 105 | 210 | 315 |
|  | All Patients |  |  |  |  |  |  |  |
| 2018-2019 season | 18-64 y | 4.09 | 70 | 34,919 | 15 | 45 | 90 | 135 |
|  |  |  | 80 | 30,555 | 17 | 52 | 103 | 155 |
|  |  |  | 90 | 27,160 | 19 | 58 | 116 | 174 |
|  | 65-74 y | 42.43 | 70 | 3,367 | 27 | 81 | 162 | 243 |
|  |  |  | 80 | 2,946 | 31 | 93 | 185 | 278 |
|  |  |  | 90 | 2,619 | 35 | 104 | 208 | 313 |
|  | 75-84 y | 107.65 | 70 | 1,327 | 35 | 106 | 212 | 317 |
|  |  |  | 80 | 1,161 | 40 | 121 | 242 | 363 |
|  |  |  | 90 | 1,032 | 45 | 136 | 272 | 408 |
|  | 75+ y | 153.30 | 70 | 932 | 72 | 216 | 431 | 647 |
|  |  |  | 80 | 815 | 82 | 246 | 493 | 739 |
|  |  |  | 90 | 725 | 92 | 277 | 554 | 832 |
|  | 85+ y | 257.56 | 70 | 555 | 36 | 109 | 218 | 327 |
|  |  |  | 80 | 485 | 42 | 125 | 249 | 374 |
|  |  |  | 90 | 431 | 47 | 140 | 280 | 421 |
|  | Patients with comorbidity |  |  |  |  |  |  |  |
|  | 18-64 y | 11.04 | 70 | 12,939 | 14 | 42 | 83 | 125 |
|  |  |  | 80 | 10,829 | 16 | 48 | 95 | 143 |
|  |  |  | 90 | 10,063 | 18 | 54 | 107 | 161 |
|  | 65-74 y | 94.28 | 70 | 1,515 | 27 | 81 | 162 | 243 |
|  |  |  | 80 | 1,856 | 31 | 93 | 185 | 278 |
|  |  |  | 90 | 1,178 | 35 | 104 | 208 | 313 |
|  | 75-84 y | 160.97 | 70 | 887 | 30 | 91 | 182 | 272 |
|  |  |  | 80 | 719 | 35 | 104 | 208 | 311 |
|  |  |  | 90 | 690 | 39 | 117 | 234 | 350 |
|  | 75+ y | 224.52 | 70 | 636 | 62 | 185 | 370 | 555 |
|  |  |  | 80 | 564 | 71 | 212 | 423 | 635 |
|  |  |  | 90 | 495 | 79 | 238 | 476 | 714 |
|  | 85+ y | 358.67 | 70 | 398 | 31 | 94 | 189 | 283 |
|  |  |  | 80 | 392 | 36 | 108 | 216 | 323 |
|  |  |  | 90 | 310 | 40 | 121 | 243 | 364 |

Abbreviations: RSV, respiratory syncytial virus; VC: Vaccine coverage

**Supplementary Figure 1.** Data sources and study periods

Post-pandemic pre-intervention period

Pre-pandemic period

| **Outcomes and database** |  | | | | | | | | | | | | |
| --- | --- | --- | --- | --- | --- | --- | --- | --- | --- | --- | --- | --- | --- |
| **HospiVir** |  |  |  |  |  |  |  |  |  |  |  |  |  |
| RSV-associated hospitalizations |  |  |  |  |  |  |  |  |  |  |  |  |  |
| **Health administrative data** |  |  |  |  |  |  |  |  |  |  |  |  |  |
| RSV-associated hospitalizations with a positive RSV test |  |  |  |  |  |  |  |  |  |  |  |  |  |
| RSV-associated emergency department (ED) visits with a positive RSV test |  |  |  |  |  |  |  |  |  |  |  |  |  |
| RSV-associated hospitalization or death after ED visit with a positive RSV test |  |  |  |  |  |  |  |  |  |  |  |  |  |
| RSV-associated death |  |  |  |  |  |  |  |  |  |  |  |  |  |
|  | 2012-13 | 2013-14 | 2014-15 | 2015-16 | 2016-17 | 2017-18 | 2018-19 | 2019-20 | 2020-21 | 2021-22 | 2022-23 | 2023-24 | 2024-25 |

HospiVir: hospital-based prospective surveillance network in Québec, Canada

| **Legend:** |  | Seasons included in the analyses |  | Intervention period |
| --- | --- | --- | --- | --- |

**Supplementary Figure 2**. Proportion of RSV and influenza identified in participants of the Québec hospital surveillance network during the surveillance periods and Quebec RSV and influenza circulation according to the provincial sentinel laboratory surveillance

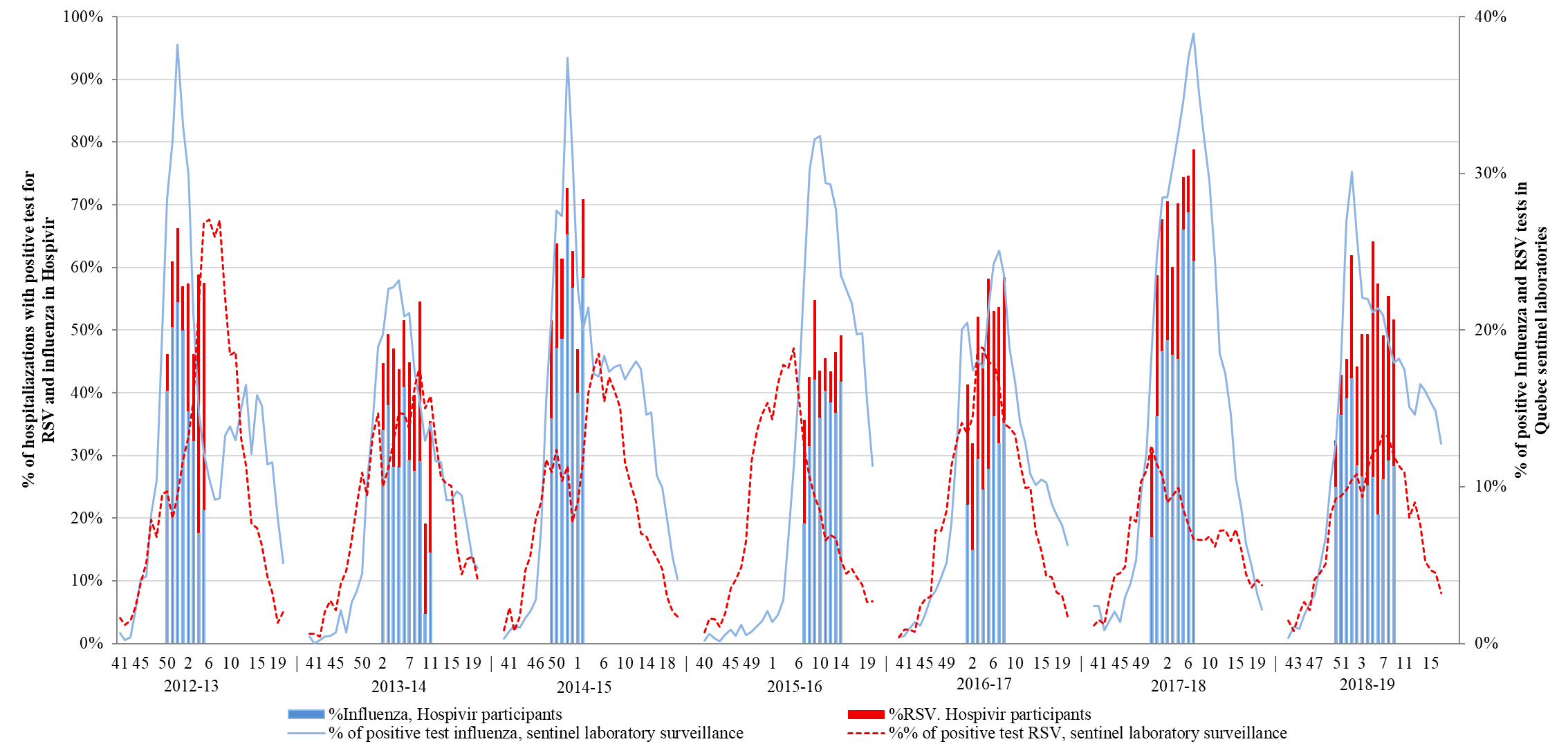

Note: Periods of the surveillance : December 9, 2012 to February 2, 2013 (epi-weeks 2012.50 to 2013.05); January 1, 2014 to March 15, 2014 (epi-weeks 2014.02 to 2014.11); November 30, 2014 to January 13, 2015 (epi-weeks 2014.49 to 2015.02); February 02, 2016 to April 04, 2016 (epi-weeks 2016.07 to 2016.14); January 1,2017 to February 25,2017 (epi-weeks 2017.01 to 2017.08); December 17, 2017 to February 17, 2018 (epi-weeks 2017.51 to 2018.07); December 09, 2018 to March 02, 2019 (epi-weeks 2018.50 to 2019.09)

**Supplementary Figure 3**. By age group, risk of emergency department visit, hospitalization or deaths among the general population aged 75 or more compared to 65-74 y (2018-19 to 2024-25 seasons), Québec, Canada.

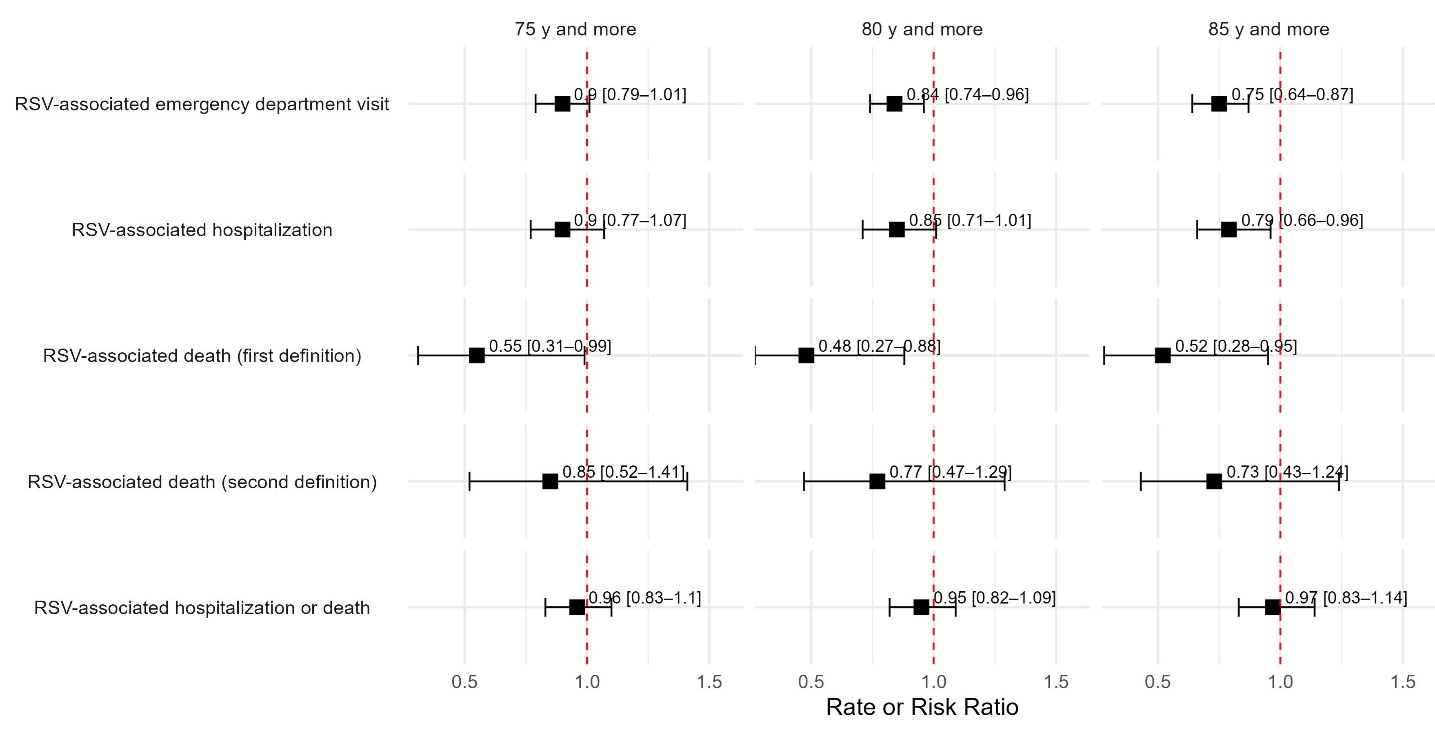

*Figure caption:* 2022-23 and 2023-24 seasons were included for RSV-associated ED visits and hospitalization, RSV-associated death cause by a respiratory disease (ICD-10 codes J00-J99; first definition) or with an RSV positive test (30 days prior to death; second definition), and RSV-associated hospitalization or death; 2018-19 to 2023-24 seasons for RSV-associated death (ICD-10 codes J12.1, J20.5, J21.0 or B97.4). RSV-associated hospitalization or death is expressed as risk ratio while other outcomes are expressed as rate ratio. The sensitivity analysis including those 80 y or more are restricted to patients with at least one comorbidity. The lateral bars refer to the 0.95 confidence interval.

**Supplementary Figure 3**. Weekly number of total RSV laboratory tests for the period from 2022-23 to 2024-25 seasons (epiweek 2022-35 to 2025-24)

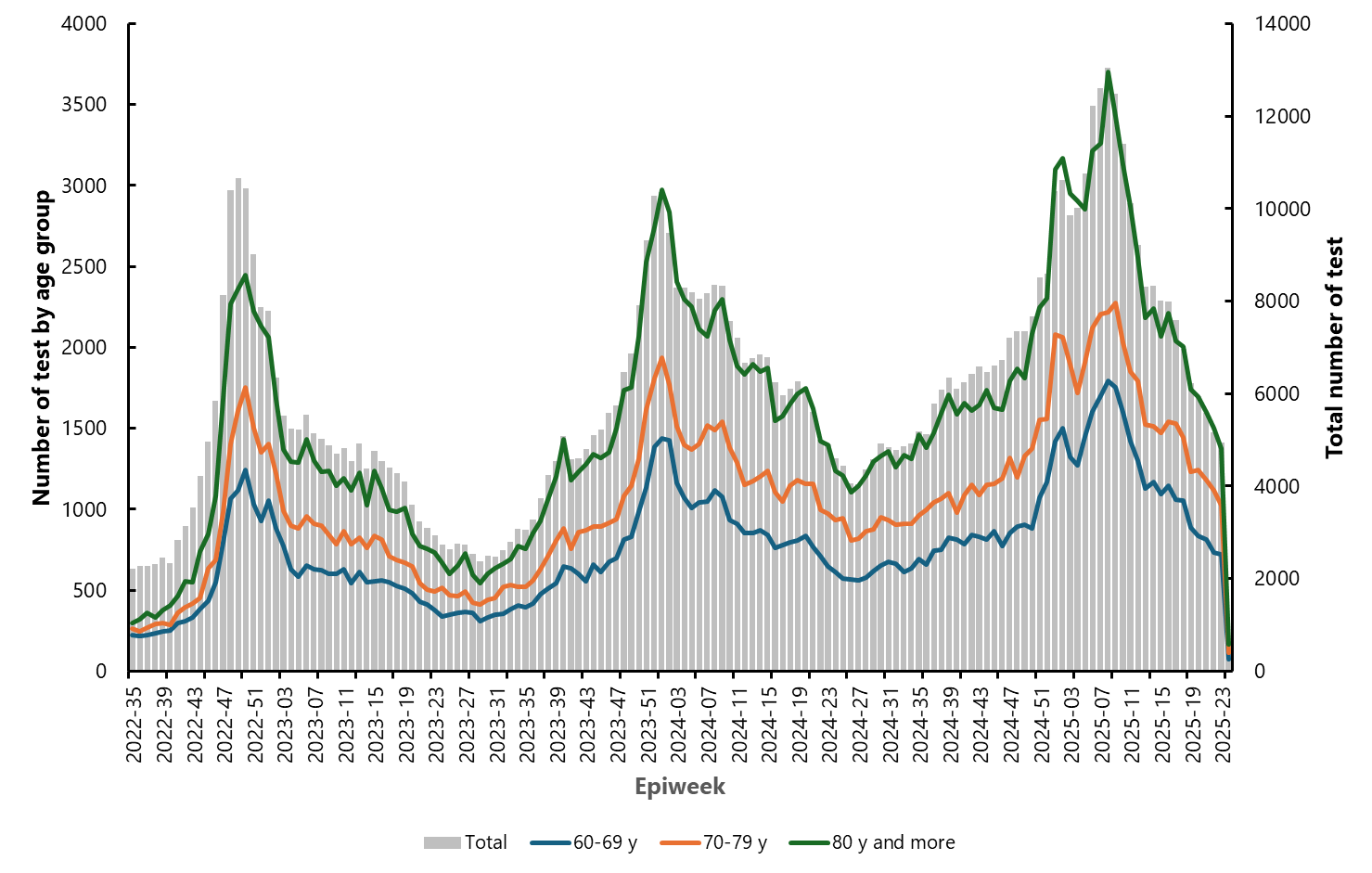

*Figure caption*: The figure shows the evolution of number of tested older adults over the past last three seasons (2022-23 to 2024-25). It demonstrates the variations in testing practices over time and the availability of tests.
